# Enhancing Cognitive Restructuring with Concurrent fMRI-guided Neurostimulation for Emotional Dysregulation: A Randomized Controlled Trial

**DOI:** 10.1101/2021.11.17.21266477

**Authors:** Andrada D. Neacsiu, Lysianne Beynel, John L. Graner, Steven T. Szabo, Lawrence G. Appelbaum, Moria J. Smoski, Kevin S. LaBar

**Author notes:** Correspondence concerning this article should be addressed to Andrada D. Neacsiu, Cognitive-Behavioral Research and Therapy Program, Duke University Medical Center (3026), 2213 Elba Street, Room 123, Durham, NC, 27710.

## Abstract

**Background:** Transdiagnostic clinical emotional dysregulation is a key component of psychopathology and offers an avenue to address multiple disorders with one transdiagnostic treatment. In the current study, we pilot a one-time intervention that combines cognitive restructuring (CR) with repetitive transcranial magnetic stimulation (rTMS), targeted using functional magnetic resonance imaging (fMRI).

**Methods:** Thirty-seven clinical adults with high emotional dysregulation were enrolled in this randomized, double-blind, placebo-controlled trial. fMRI was collected as participants were reminded of lifetime stressors and asked to downregulate their distress using CR tactics. fMRI BOLD data were analyzed to identify the cluster of voxels within the left dorsolateral prefrontal cortex (dlPFC) with the highest activation when participants attempted to downregulate, versus passively remember, distressing memories. Participants underwent active or sham rTMS (10 Hz) over the target while practicing CR following autobiographical emotional induction.

**Results:** Receiving active versus sham rTMS led to significantly higher high frequency heart rate variability during regulation, lower regulation duration, and higher likelihood to use CR during the week following the intervention. There were no differences between conditions when administering neurostimulation without CR compared to sham. Participants in the sham versus active condition experienced less distress the week after the intervention. There were no differences between conditions at the one-month follow up.

**Conclusion:** This study demonstrated that combining active rTMS with emotion regulation training significantly enhances emotion regulation and augments the impact of training for as long as a week. These findings are a promising step towards a combined intervention for transdiagnostic emotion dysregulation.

---

Emotional dysregulation is a key problem that cuts across psychiatric disorders and is associated with considerable burden of illness. Conventional approaches to psychiatric treatment emphasize remediating neurobehavioral processes associated with the downregulation of emotional arousal. For instance, emotion regulation skills building, a component of many cognitive behavioral therapies, is effective in decreasing psychopathology across affective disorders(1, 2); nevertheless, selective treatments that target transdiagnostic emotional dysregulation are still lacking. Improvement in emotional functioning happens at a slow pace and is limited in magnitude and scope(1, 3). Therefore, there is a critical need to develop new therapeutic approaches that integrate current knowledge about the neurophysiology of emotion regulation into clinical practice. Specifically, combined treatments that manipulate neurobiological pathways of emotion regulation concurrently with psychotherapy could increase training efficacy by facilitating learning, improving compliance by reducing therapy duration, and expanding functional gains beyond what traditional psychotherapies can achieve alone(4).

Emerging neuroimaging findings highlight several disruptions in brain function that are associated with clinical emotion dysregulation. When performing emotion regulation tasks, hypoactivation in prefrontal regions such as the dorsolateral (dlPFC), ventrolateral (vlPFC)(5), and dorsomedial prefrontal cortex (dmPFC)(6), and hyperactivation in limbic/paralimbic brain areas, such as the left anterior insula(6) and amygdala(5), characterize difficulties in changing emotional arousal in anxiety and mood disorders. Importantly, the tasks used for these neuroimaging studies overlap with skills taught in cognitive behavioral therapy (CBT), especially with cognitive restructuring (CR). Meta-analytic studies of CR have consistently identified activation in this emotion regulation network in healthy subjects(7) and in anxious and depressed adults(6). Although this fronto-limbic network may not encompass all modes of emotion regulation (e.g., meditation practice extends to additional brain regions)(8), these findings provide a rigorous scientific foundation for hypothesizing a neural signature of transdiagnostic emotion dysregulation in response to negative arousal(9).

Thus, a transdiagnostic intervention for emotional dysregulation can be constructed to alter this neural dysfunction. CR is a fundamental skill taught in many evidence-based psychotherapies for affective disorders(10-12) that has also been studied in basic neuroscience(13, 14). CR involves thinking differently about a situation in order to change the emotional experience and is currently one of the most studied emotion regulation strategies across both basic and clinical research(15). Effective use of reappraisal during an emotion regulation task increases high frequency heart rate variability (HF-HRV)(16), a marker of effective emotion regulation(17, 18). Functional neuroimaging studies examining CR have found activation distributed throughout a fronto-limbic network, including the dlPFC, vlPFC, ACC, superior frontal gyrus (SFG)/frontal pole, amygdala, and insula(6, 13, 14). Theoretical and empirical findings highlight that CR can be further subdivided into specific tactics, such as reframing and distancing (6, 13), although these tactics largely overlap in their neural representation.

In this study, we propose to augment CR practice with neurostimulation targeted towards the same regulatory neural network in order to shorten the required time to learn this skill. Emerging evidence suggests that noninvasive neurostimulation, such as repetitive transcranial magnetic stimulation (rTMS) can alter emotional processes(19) and enhance performance on affective tasks(20) in healthy samples, especially stimulation is administered during an active task(21). In transdiagnostic clinical adults, active versus sham rTMS applied over the left and right dlPFC conducted in conjunction with CR practice led to significant enhancement of physiological (HF-HRV), behavioral (regulation duration), and self-report (subjective distress) indices of emotion regulation, up to a month later in a transdiagnostic clinical sample(22). The hypothesized mechanism is that neurostimulation facilitates neural processing in localized cortical regions via inducing Hebbian-like plasticity(23) which may have therapeutic effects and may remediate deficits in affected neural networks(24).

Increases in HF-HRV during emotional arousal are correlated with and may be a sensitive index of enhanced emotion regulation(25-27). Participants who engage in effective emotion regulation have higher changes in HRV from baseline than those who do not engage in regulation(17, 28). Also, successful increase in emotion (via exaggeration) during positive and negatively arousing film clips was strongly connected with change in HRV from baseline(29). A meta-analysis also found a strong connection between HF-HRV and use of cognitive strategies to manage emotions(30). Furthermore, increases in HRV are correlated with the amount of constructive coping adults report they use when distressed(25, 31) as well as with successful pursuit of goal under emotional arousal(32, 33). Taken together, these findings strongly suggest that HF-HRV can be a reliable(27) marker of effective emotional regulation in the context of emotional arousal.

The current double-blinded, placebo-controlled trial extends the prior literature by using individualized fMRI-guided rTMS to optimize the effect of the rTMS-CR intervention on emotion regulation in transdiagnostic adults. This approach ensures that inter-individual differences in brain anatomy and physiology that are crucial for fine-grained targeting are fully accounted for, optimizing our ability to harness the full potential of neurostimulation technology(34). Prior findings suggest that using an individualized fMRI stimulation target could yield better rTMS outcomes(35, 36). We hypothesized that when compared to sham rTMS, active rTMS administered in conjunction with CR will enhance emotion regulation in the moment and up to a week after the intervention. In addition to our primary outcome measures, we planned to explore long-term effects of this intervention and differences in response based on level of proficiency with CR at the beginning of treatment.

## Materials and Methods

### Participants and Procedures

This study was an adjunctive supplement to a previously reported examination of rTMS enhanced CR(22). Both studies were pre-registered under the same Clinical Trials ID (NCT02573246) and ran concurrently between April 2016 and February 2020. The results included here are from the adjunctive supplement only. Both studies were powered using a one-time comparison between sham and active neurostimulation in potentiating emotion regulation in healthy adults (Cohen’s *d* effect sizes = .94 - 1.87 for physiological and self-report differences in emotion regulation between conditions)(20). A G-Power analysis(37) indicated that 15 participants would be needed to observe an effect size of .94. Therefore, we intended to treat 20 participants per condition.

Participants were recruited through online websites (e.g., Craigslist), flyers, and physician referrals. Interested participants completed an online and in-person screen before being invited for an imaging session. Of 46 treatment-seeking community participants who were eligible, 31 participants presented to the intervention session and are considered the intent to treat (ITT) sample. ITT participants were 6 men and 25 women between the ages of 18 and 62 (M = 33.77; SD = 13.48 years old) who met criteria for any DSM-5 disorder (excluding active substance use, psychotic disorders, and Bipolar I) and who self-reported above average emotional dysregulation. Participants met criteria for an average of 3.29 (*SD =* 1.68) current diagnoses and 4.94 (*SD* = 2.11) lifetime diagnoses according to the structured interview for *DSM-5* disorders (SCID-5)(38). The majority of participants (*n* = 28) met criteria for an anxiety disorder, over half met criteria for a current depressive disorders (*n* = 19), 12 for a compulsive disorder, 5 for an eating disorder, and 13 for a stress disorder (see further breakdown in Supplement). Eight participants (25.80%) met criteria for at least one personality disorder according to the SCID-5-PD(39).

High emotional dysregulation was operationally defined as a total score of 89 or higher on the Difficulties in Emotion Regulation Scale (DERS; additional description in Supplement)(40). ITT participants scored on average 109.55 (SD = 16.59) on the DERS, The sample was further stratified using the Emotion Regulation Questionnaire (ERQ)(41) cognitive reappraisal subscale (range: 1-7). We aimed to include in this study adults with a varied proficiency in using CR. Therefore, we established cutoffs for low, moderate and high reappraisal proficiency and randomized participants in conditions accounting for this variable (details for how cutoffs were computed are provided in the Supplement). ITT participants scored an average of 4.24 (SD = 1.25) on the ERQ reappraisal subscale. The breakdown in CR proficiency between groups is presented in Supplementary Table 1. Additional exclusion criteria are presented in the Supplement (see CONSoRT **Figure 1** for details). Participants received a maximum compensation of $250. The study was approved by the Duke University Health System Institutional Review Board, and it involved a total of five sessions and a week of ambulatory data collection (**Figure 2A**).

**Figure 1.**
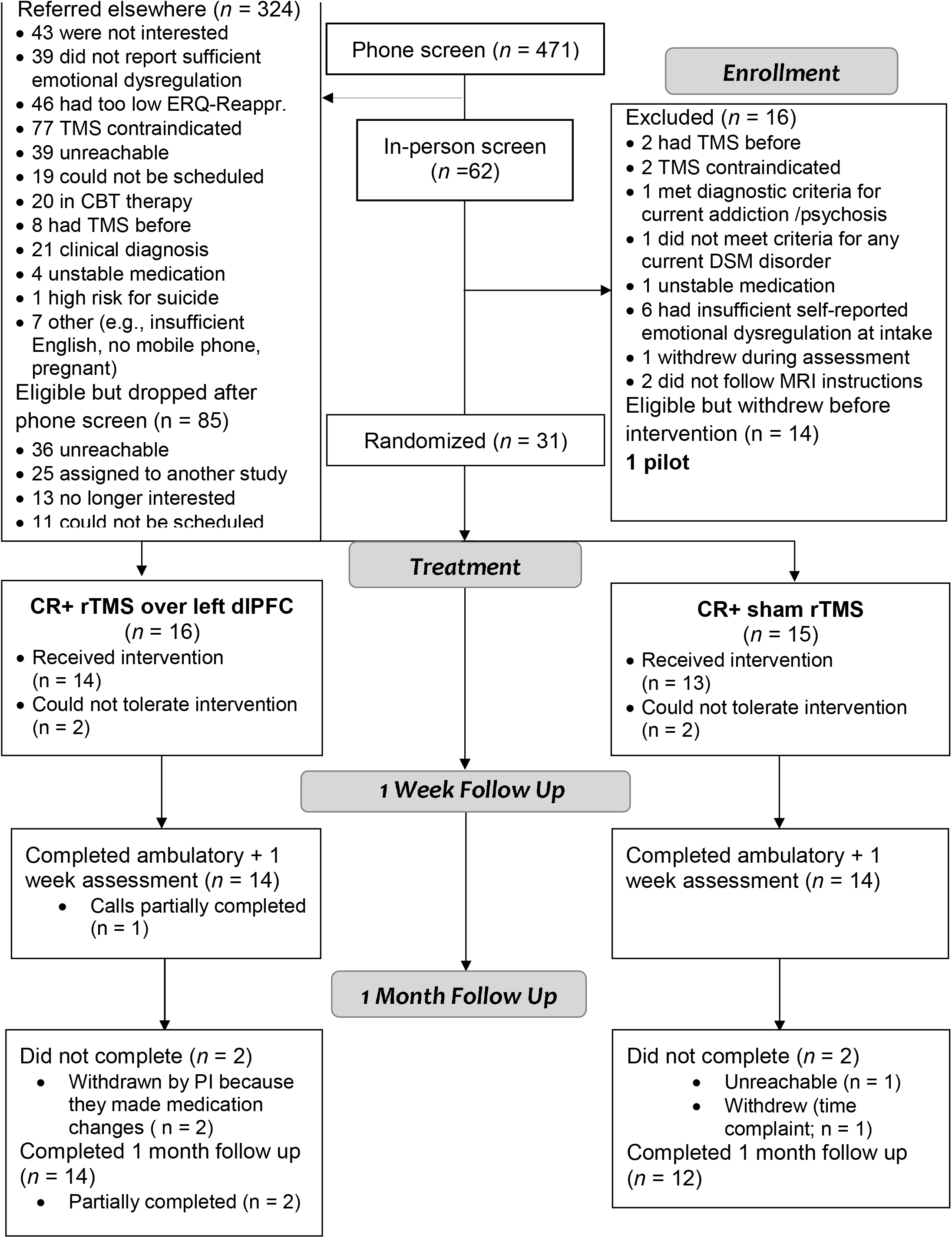
Participant enrollment, treatment assignment, and attrition. CR = Cognitive restructuring; rTMS = repetitive transcranial magnetic stimulation; ITT = Intent to Treat; dlPFC = dorsolateral prefrontal cortex

**Figure 2.**
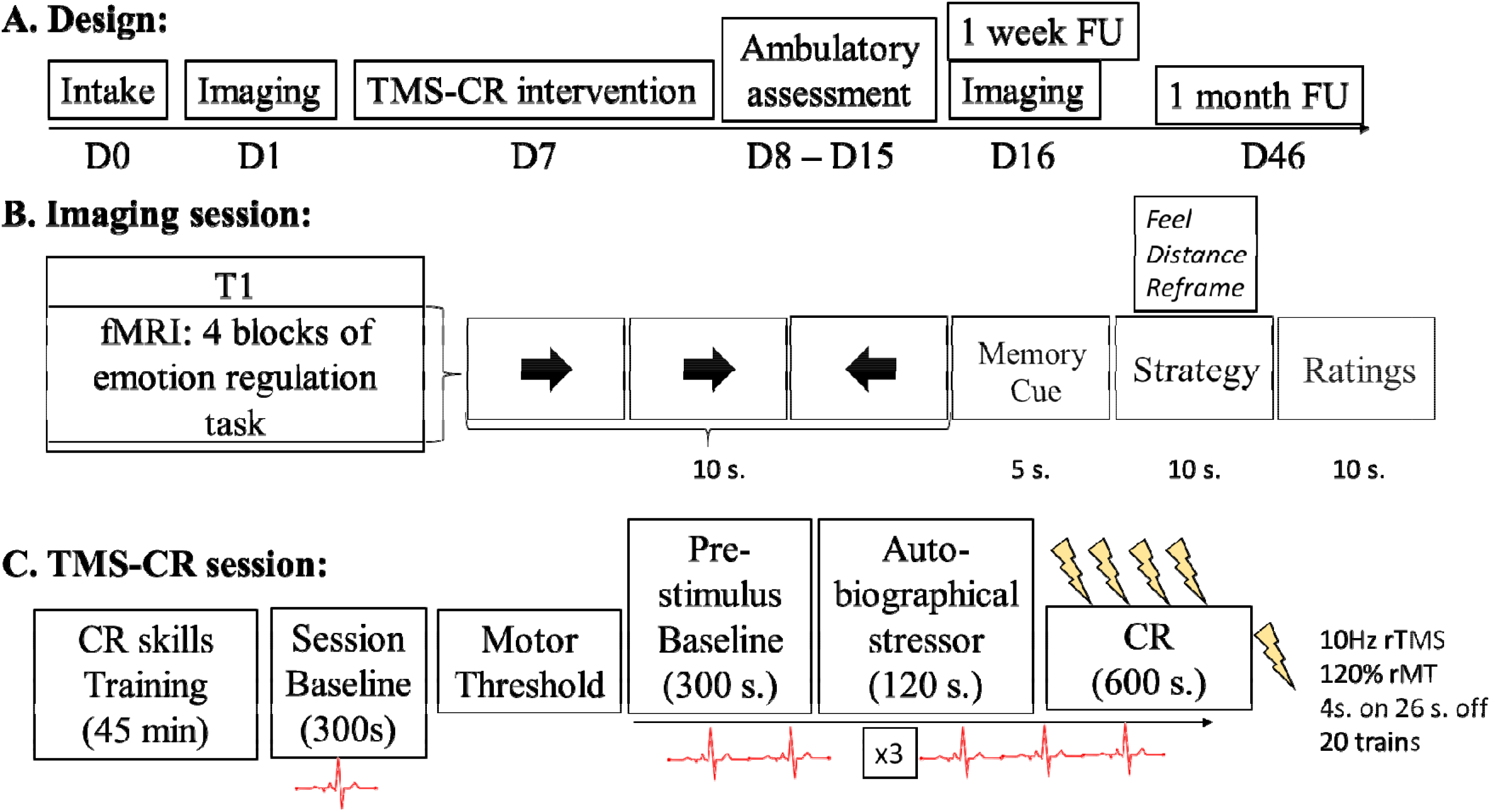
A. Overall study design (with target days since intake when each session was planned to occur), B. Schematic of the imaging session including a description of the task (cues are either for negative or neutral memories), C. Schematic of the TMS-CR intervention. TMS = repetitive transcranial magnetic stimulation (either active or sham); CR = cognitive restructuring.

#### Intake Session

After providing voluntary, written informed consent, participants completed diagnostic assessments (SCID-5, SCID-PD), a verbal intelligence test(42) and a questionnaire packet including the ERQ(41), DERS(40), the Work and Social Adjustment Scale (WSAS)(43), and the Outcome Questionnaire-45 (OQ-45; see Supplement for psychometric information)(44). Four autobiographical negative emotion inductions were developed with each participant using established methods(45, 46) (see Supplement). These personalized scripts were narrated and digitally recorded into .wav audio files to be presented in random order during the intervention and follow up sessions.

Participants also identified 10 negative and 4 neutral lifetime memories, rated the arousal and valence remembering this memory caused them(47). Each memory was rated along several phenomenological characteristics (e.g., arousal, vividness) using an established questionnaire(48, 49). Participants then generated a unique verbal descriptor for each memory to be used as a retrieval cue during fMRI(48). Together with the four autobiographical memories, participants provided 14 recent and distal negative memories.

#### Imaging sessions

As soon as possible after intake, and a week after the rTMS+CR intervention, participants were asked to complete an MRI session. These sessions were identical in order to assess pre-post differences from our intervention. In this paper, only data from the pre-treatment session are used to describe the TMS targeting procedure. During the first hour, participants were trained in the emotion regulation task following established procedures (see details in Supplement)(47, 50). Participants were reminded of their memory cues, and practiced identifying the memory by just seeing the cue word until all cues successfully triggered autobiographical memories. In addition, participants were introduced to the *distancing, reframing*, and *feel* strategies. Participants were taught that *reframing* involves looking at a situation from a different perspective, or thinking of what might come out of the situation in order to feel less upset. For *distancing*, we instructed participants to either look at the situation objectively, like a news reporter, think that the situation happened in a different space, or think that it’s a time limited event or that it’s just a memory. Last, we asked participants to let their thoughts and emotions flow naturally without expending efforts to change experience when being prompted to *feel*.

Next, participants moved into the scanner (**Figure 2B**). The scan started with an anatomical image (3-D T1-weighted echo-planar sequence, acquisition matrix = 256 × 256, time repetition [TR] = 2300 ms, echo time [TE] = 3.2 ms, field of view [FOV] = 256 mm^2^, in-plane voxel size = 1.0 × 1.0 mm, slice thickness = 1.0 mm, spacing between slices = 0 mm, 162 slices). Four runs of EPI functional images were then acquired with an oblique axial orientation (acquisition matrix = 128 × 128, TR = 2000 ms, TE = 30 ms, FOV = 256 × 256 mm^2^, in-plane voxel size = 2.0 × 2.0 mm, slice thickness = 2.0 mm, spacing between slices = 0 mm). Runs 1 and 3 had 240 volumes; runs 2 and 4 had 216 volumes.

During the four runs of functional acquisitions, participants performed 40 trials of the emotion regulation task. Each trial of the task consisted of sequential presentations of: a jittered cross (passive baseline), an arrow task (active baseline), a memory cue word (generated at the intake session; negative/neutral), a strategy word (“Feel”, “Distance”, or “Reframe”), and a rating of distress after the regulation attempt (range: 0-9; see Figure 2 B). Behavioral responses were recorded with a 4-key fiber-optic response box (Resonance Technology, Inc.). Data from the first imaging session were used to develop TMS targets. Data from the follow up imaging session are presented elsewhere.

#### Randomization

Following the first MRI visit, 31 ITT participants were randomly assigned to active rTMS stimulation (*n* = 16), or sham stimulation (*n* = 15). Participants were matched according to sex at birth (male/female), use of psychotropic medication (yes or no), and proficiency with reappraisal (low/moderate/high) using a minimization randomization algorithm(51, 52). Two sealed envelopes were created for each participant, one to be opened by the TMS technician who set up the coil before the intervention for either the sham or active paradigm, and one to be opened at the end of the study by the assessor and participant to discuss randomization. The subject and the PI who conducted the intervention were kept blind until the end of the intervention.

#### Neurostimulation target identification

FMRI analyses were performed after the first MRI session to define the individualized stimulation target. Structural and functional data were first examined with MRIQC and preprocessed with fMRIprep1.1.1(53) (see Supplement). fMRI data were then skull-stripped(54) and underwent high-pass temporal filtering with a 100-second cutoff. At the first level, functional data were analyzed as individual runs, using a general linear model (GLM) as implemented in FSL’s FEAT procedure(55). Model regressors were created for the arrow task (9 s), memory cues (5 s), “distance” strategy (10 s), “reframe” strategy (10 s), “feel” strategy after the negative memory (“Feel Negative”; 10 s), “feel” strategy word after neutral memory (“Feel Neutral”; 10 s), and distress ratings (10 s). A weight of 1 was attributed to each task regressor in the general linear model (GLM). Trial “on” times were convolved with a double-gamma hemodynamic response function to create the final GLM regressors. The “Restructure vs. Feel Negative” contrast was defined as the combination of distancing and reframing minus the “Feel Negative” instructions. This contrast was then used in the second-level analysis, in which the first-level results for each of the four runs were combined using a fixed-effect model. The statistical map of the “Restructure vs. Feel Negative” contrast was transformed to native space back from standard space using the inverse ANTS transformation(56), and overlaid onto the anatomical image in a neuronavigation software (BrainSight, Rogue Research, Canada). The cluster within the left dlPFC showing the strongest positive z-value for the contrast was defined as the target. In order to be selected, the cluster had to be visible on the surface of the rendered brain in the BrainSight software. The coil orientation was visually set up so that the induced E-field was perpendicular to the closest sulcal wall. When this orientation was not feasible (e.g., coil handle in front of participant’s face), the symmetric orientation was used, and the current was reversed. Scalp-to-cortex distance in mm at the site of stimulation was measured using the tape tool in BrainSight. See **Figure 3** for a visual depiction of all personalized targets and specific examples.

**Figure 3.**
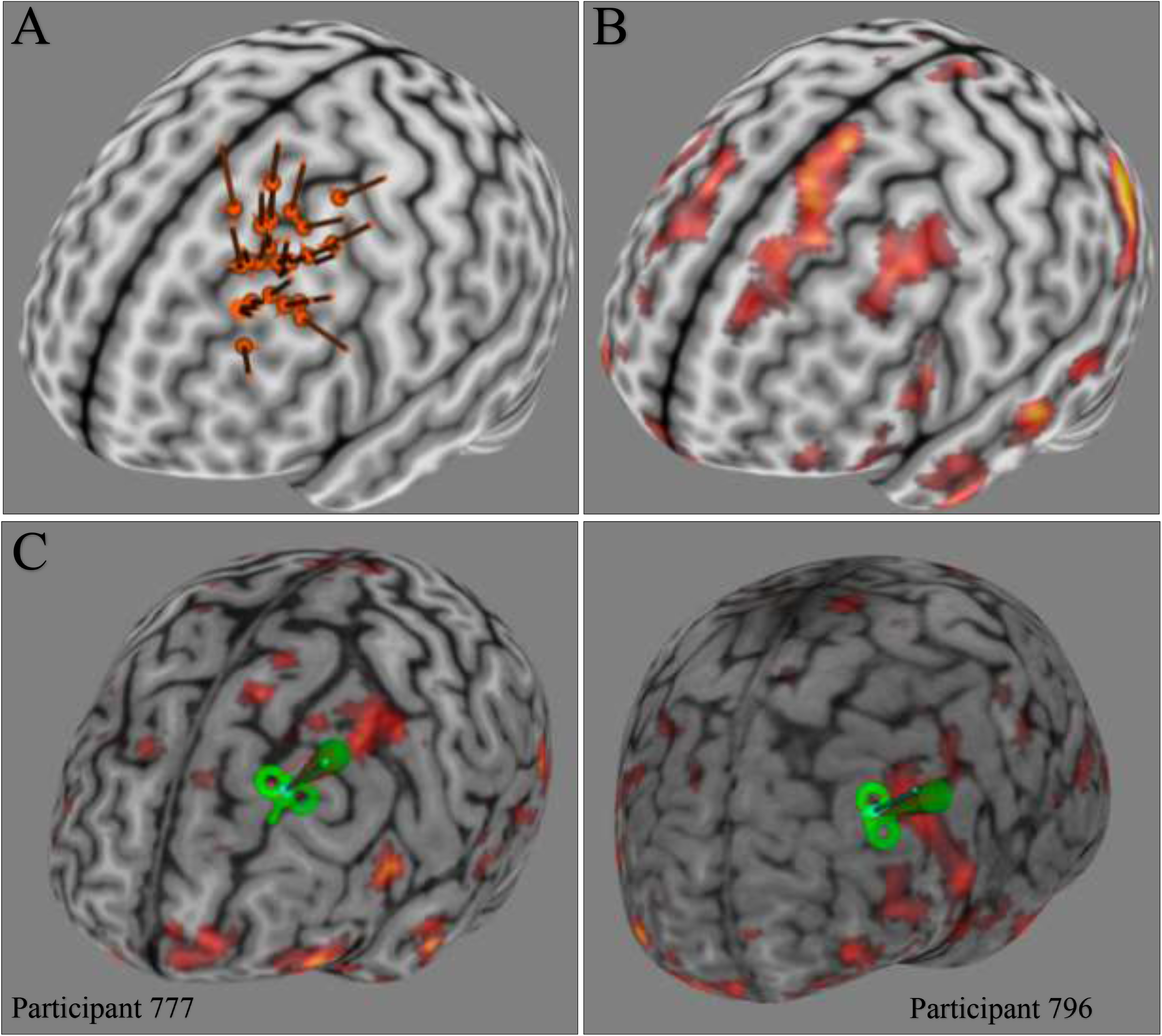
**(A)** All the personalized targets for TMS based on the highest difference in activation between restructure and feel negative instructions during the first MRI session transformed from native space into MNI space. These targets corresponded to the activation map at the individual level, and were then translated into MNI space for visualization (**B)** Group average activation map for contrast of interest [restructure – feel negative] generated using a mixed effects (FSL’s FLAME 1) whole-brain analysis using cluster correction following a voxel-wise Z-score threshold of 1.96 show significant activation clusters in the left dlPFC, the region selected for personalized targeting (**C**) Examples of personalized targets from two participants.

**Figure 4.**
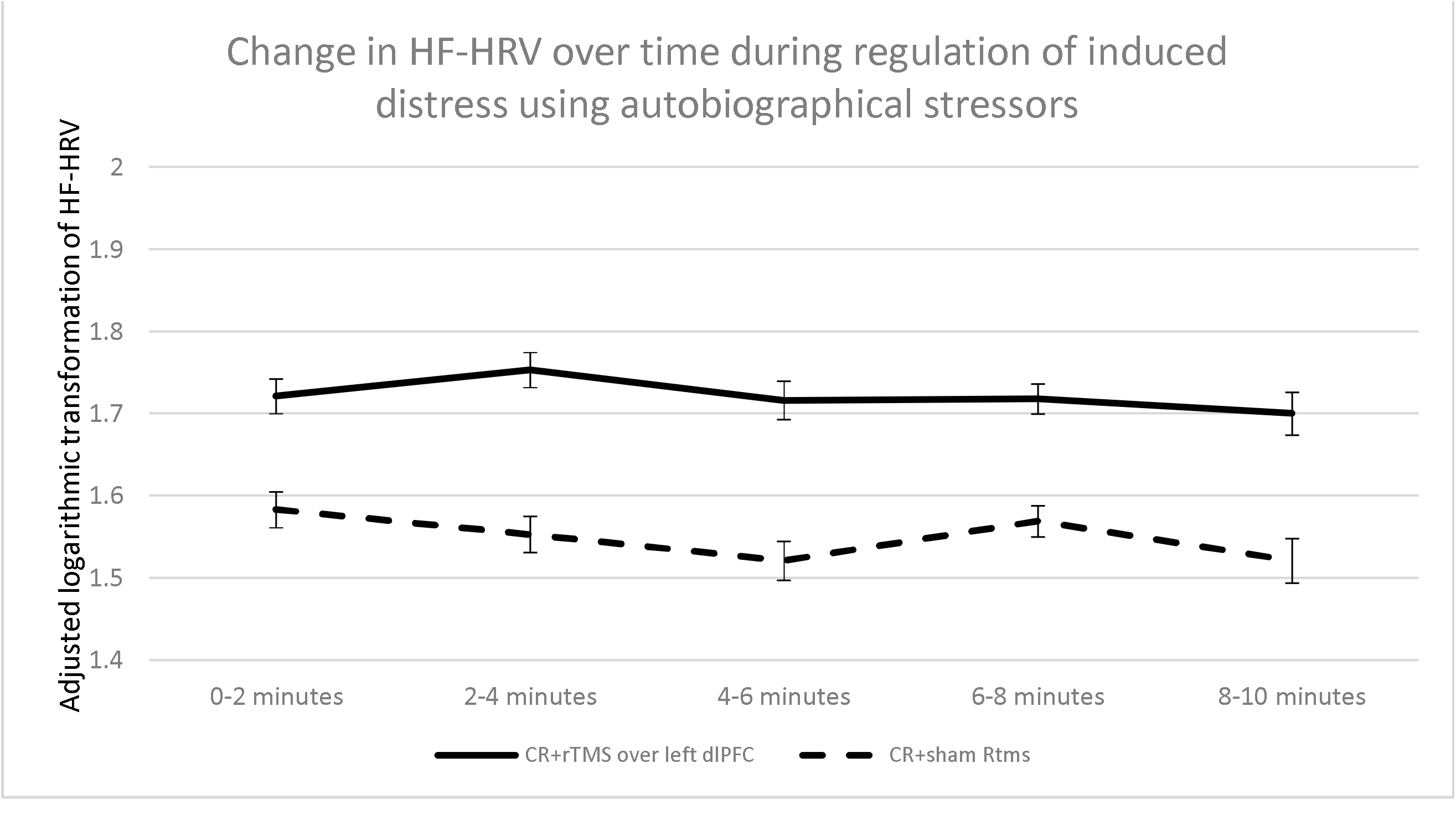
Changes in high frequency heart rate variability (HF-HRV; a marker of effective emotional regulation) summarized across the three regulation periods allotted during the intervention, and separated by condition, and by time segment. The original HF-HRV value was measured in seconds^2^, multiplied by 100000 and transformed using a log function to achieve normality. Each time point on the graph represents the estimate marginal mean from the LSD pairwise comparisons from the MMANOVA main analysis which accounts for covariates. Regulation periods followed autobiographical negative emotional inductions.

#### Intervention Session

Participants returned for the 3.5-hour intervention session as soon as possible after the target was identified and no later than month since intake (**Figure 2C**). The first 45 minutes were spent on skills training, one-on-one with a trained clinical psychologist, and was focused on learning CR and practicing on standardized and personal examples. Skills training used standardized procedures blending psychotherapeutic approaches(57, 58) with instructions in CR that matched prior neuroimaging studies(59). Participants were taught distancing(13, 60) and reframing(57, 59, 61, 62), practiced on standardized and personal examples, and had to pass a quiz to ensure they correctly understood the procedure (see Supplement).

Following successful completion of the behavioral training, electrodes recording heart rate (HR) and skin conductance were placed on the participant’s wrist, ankle, and fingers. Amplified analog data were converted to digital recording and filtered using BIOPAC’s AcqKnowledge 4.1 software. Psychophysiological measurements were collected for a 300 s session baseline and then continuously during the combined intervention using the BIOPAC MP150 recording system (Goleta, CA).

##### rTMS Intervention

Active or sham rTMS was performed with a figure-8 coil (A/P Cool-B65) and a MagPro X100 stimulator (MagVenture, Denmark) set up to deliver biphasic pulses. 10 Hz rTMS was performed following standard FDA-approved parameters(63, 64): 4 s of stimulation and 26 s of an inter-train interval at 120% rMT. Neurostimulation was administered for four discrete 600-s periods, which included 20 trains each, for a total of 80 trains for the session. This approach has been found to be effective in modulating emotion regulation in our previous study(22). Sham stimulation was applied using the same intensity setting but with the coil in placebo mode, which produced similar clicking sounds and somatosensory sensations (via electrical stimulation with scalp electrodes) without a significant magnetic field reaching the brain(65). This type of sham stimulation allowed participants to stay blinded to the type of stimulation they received, as reflected in the results section.

For each subject, stimulation was delivered over the personalized left dlPFC target. Coil position and orientation were continually monitored through a stereotaxic neuronavigation system (Brainsight, Rogue Research, Canada). Each TMS session was conducted by the principal investigator (PI; first author) with the assistance of a TMS technician. The PI, who was blinded to the stimulation condition, led the participant through the session, decided on dose adjustments and course of action for any protocol deviations. For example, if a participant could not tolerate rTMS, the PI could decide to drop the intensity to rMT and slowly ramp it back up during habituation. The TMS technician was not blinded to the condition and prepared the coil but did not influence the course of the session or interact with the participant. Given the MacVenture sham set up, adjusting intensity of stimulation did not break the experimenter’s blind.

The TMS-CR intervention started with a 600-s habituation period during which active or sham rTMS was applied while participants were not given any instructions. Then, the intervention proceeded with a task that repeated three times with breaks in between. The task consisted of a 300-s pre-stimulus baseline, hearing one of the personalized stressors developed at the intake session and being instructed to imagine it as vividly as possible (120s), and engaging in emotion regulation using CR (600s) while concurrently receiving active/sham neurostimulation. After each baseline, stress induction, and regulation period the participant was asked to rate subjective units of distress (SUDS) and dissociation(66). At the end of the intervention, participants guessed whether they received real or sham rTMS and rated side effects.

#### Ambulatory Assessment and Follow-Up Visit

Starting the day after the intervention, participants received 8 calls per day for 7 days at pseudo-random times. On each call participants were asked if they had used CR since the previous call (yes/no) and what was their current distress (0-no distress at all to 9-extreme distress). At the end of the week the battery of self-reports from intake was administered again via an online link and the follow up MRI was conducted.

Participants returned to the research office a month later to complete a stressor task without neurostimulation, an acceptability and feasibility interview, and the battery of self-reports for a third time. The stressor task included measuring HR and skin conductance while the participant stood still for a 300-s baseline, underwent a 120-s stressor induction using a fourth autobiographical stressor, and engaged in prompted CR for 300 s. SUDS were rated after each experimental task.

An unpublished, previously developed in-house interview (22) was then used to examine feasibility and acceptability as directly relevant to the study (see Supplement) rated on a scale from 0 (not at all) to 9 (extremely). Satisfaction was rated on a 0 (low) to 100 (high) continuous scale. Two participants had to discontinue the study early, and therefore participated in the exit interview at the 1-week follow up. After the exit interview, the blind was broken, and the experimenter and the subject were debriefed about the study upon knowing the assigned condition.

#### Psychophysiological Data

HR was recorded during the whole duration of the intervention session and the follow-up session as a measure of emotion regulation. AcqKnowledge software was used to collect and pre-process raw ECG data by using a built in automated HRV analysis tool that follows established frequency domain algorithm guidelines(26). The regulation period was divided into 120-s bins, and HF-HRV was extracted from each bin. HF-HRV values were extracted from the last 120 s of each baseline, from each stressor period, and from the five bins in each regulation period and habituation (see additional details in the Supplement). HR was calculated as beats per minute using the ‘Find Rate’ function of AcqKnowledge with a moving average with a window of 15 seconds. Average baseline was computed from the last 240 s of each pre-stimulus baseline.

*Regulation duration* was defined for each regulation period as the amount of time it took from the onset of regulation instructions for the continuously monitored HR to reach a value that was lower or equal to the average pre-stimulus baseline HR. If the person started the regulation period below the average pre-stimulus baseline, we coded the regulation duration as “never stressed” and did not include it in analyses. If the participant never returned to baseline in the 600 s allocated to the regulation period, we set the regulation duration time to be 600 s. To create a baseline for this variable we measured the time it took to return to baseline during habituation following the onset of neurostimulation. At follow up, regulation duration was defined identically, except there was no baseline value computed, and the maximum value the variable could take was 300 s.

### Statistical Analyses

All analyses compared the effect of sham to active rTMS over the left dlPFC. Preliminary analyses, including t-tests and chi squares, were conducted to assess demographic differences between treatment conditions. We also examined any differences on randomization variables (gender, use of medication) and other potential confounding variables like dissociation during regulation, or arousal induced by rTMS alone. Planned covariates included baseline measurements of the outcomes and scalp-to-cortex distance(67, 68) for outcomes measured during intervention and the ambulatory week. Scalp-to-cortex distance was included as a covariate given evidence that higher distance may impede the effectiveness of neurostimulation interventions for emotion regulation that are not designed to account for this parameter(69).

Primary outcomes set *a priori* were regulation duration and HF-HRV during regulation. Secondary outcomes included self-reports (SUDS during intervention, SUDS and self-reported use of CR during the ambulatory week, and DERS, OQ-45, ERQ-Reappraisal, and WSAS at follow up). Mixed-effects hierarchical linear models (MMANOVA) with analytically determined covariance structures were used to analyze the repeated measures data(70). All MMANOVA models used a restricted estimated maximum likelihood model to account for missing data(71)(i.e., cases with missing data were not discarded, but slopes for each participant were computed with the data available). Estimated marginal means (EMMs) were compared using LSD corrections for significant main and interaction effects, and are presented in the results section. Significance values from primary and secondary outcome main effects and planned covariate effects were ranked, and a Benjamini and Hochberg false discovery rate (FDR) correction (72)(*q* = 0.05) was employed to account for multiple comparisons.

To test immediate effects of the intervention, we conducted three analyses examining HF-HRV, SUDS, and regulation duration. The treatment condition (active or sham rTMS), the experimental condition (regulation 1, 2, and 3), the time within each experimental condition (coded 0-4 for each 120s segment within that period), and both session and pre-stimulus baselines were used to predict HF-HRV. Treatment condition, experimental condition, and experimental period (post-baseline, post-post-stressor, post regulation), and planned covariates were used to predict intervention day SUDS. For regulation duration, baseline, treatment condition, and experimental condition were included as fixed effects. Effect sizes were computed by using Feingold’s formula(73) and interpreted using Cohen’s specifications(74). Time was considered a categorical variable.

To test near-term effects of the TMS-CR intervention a hierarchical linear models (HLM)(75) was used to examine condition differences in SUDS during the ambulatory assessment using the SUDS rating at the beginning of the intervention day (before any procedures were conducted) as baseline and scalp to cortex distance as a covariate. To examine use of CR (indicated as yes/no on each call) we utilized a Generalized Estimating Equations (GEE) binary logistic model (logit link, unstructured working correlation with robust estimators)(76, 77). Intake ERQ reappraisal and scalp-to-cortex distance were included as a co-variates. Call days were considered a continuous time variable, and call numbers (first, second, …) within the day were coded categorically.

To test the long-term effects of the intervention, six MMANOVA models were conducted: four examined between-treatment condition differences at the one-week and one-month follow up (one for each of the ERQ, DERS, OQ-45, and WSAS measures), and two additional models examined HF-HRV and SUDS, respectively, during the follow-up stressor task. An independent samples *t*-test examined differences in regulation duration at follow up. For the longitudinal self-reports (DERS, OQ-45, WSAS, and ERQ Reappraisal), time was entered as a categorical variable and intake measurements were co-varied. Differences between levels of proficiency with CR at intake were examined across all outcomes in exploratory analyses that were not FDR corrected.

## Results

### Missing Data

Out of 31 ITT participants, one felt faint during the MT procedure and stopped all study procedures at that point. Three participants found rTMS too painful and withdrew during the habituation portion of the intervention (one in sham and 2 in active conditions). Therefore, 27 participants completed the intervention session and follow up assessments (see supplementary Table 2 for demographic data). All ITT available data were included in the outcome analyses. Across the psychophysiological and self-report outcome measures, only 5.06% of the data was missing (see details in the Supplement). Two participants had limited follow-up data because they changed medications and were withdrawn from the study. Of the 1512 calls placed during the ambulatory assessment, participants provided data on 1128 (74.6%) of the calls.

### Normality Assumption

All variables were tested to examine whether they violated the normality of distribution assumption using Shapiro-Wilk normality tests (*W* < .90). At both intervention and follow up, HF-HRV was transformed to normal using the function lg10 (HF-HRV*1000000). The regulation duration variable was closest to normality when transformed using a lg10 function. All other variables were normally distributed.

### Preliminary Analyses

There were no significant differences between conditions in age, gender distribution, marital status, sexual orientation, ethnicity and race, income, self-reported use of reappraisal and emotional dysregulation, use of psychotropic medication, or number of current or lifetime diagnoses met (*p*s > .05). Therefore, our randomization procedures were successful, and none of these variables were covaried.

During experimental procedures, self-reported dissociation was minimal (M = 0.31, SD = 0.98; Range 0-10) and not different between conditions. In addition, there was no significant difference between treatment conditions in lg_HF-HRV (*F*_treatment_condition_ [1, 22.93] = 1.16, *p* = .29; M_lg_HF-HRV_ = 1.71; SD = .40) or SUDS (*t*[25] = .67, *p* = .51; M_SUDS_ = 1.7; SD = 1.79) during habituation. Therefore, these variables were not included as covariates. The SD for HF-HRV during habituation and for SUDS post habituation were used to compute effect sizes.

### Tolerability, Acceptability, and Blinding Quality

On average participants were 71.44% (SD = 20.90) likely to recommend this treatment to a friend (reported range 21% – 100%). Participants found the overall study procedures very acceptable (M_Acceptability_ = 7.28, S.D. = 1.14) and feasible (M_Feasibility_ = 6.41, S.D. = 0.90) with no differences between conditions (*p*s > .05). When asked the open-ended question “what was it like being in our study?”, participants reported finding the study very interesting and the intervention useful. Participants observed that feasibility would be improved with fewer phone calls and shorter visits. Two participants mentioned rTMS being uncomfortable. Five participants in the active condition (35.7%) and three in the sham condition (23.0%) reported worsened headache, and 11 participants (6 in active, 5 in sham) reported worsening neck pain after the intervention. One person in active and three in sham reported a reduction in headache immediately after the intervention. All headache and neck discomfort improved 2 days after the intervention. One participant described a hypomanic episode starting the day after the neurostimulation procedure that she addressed with a medication change leading to withdrawal from the study.

A chi square analysis indicated no significant difference between treatment conditions in guessing assignment to active/sham (χ^*2*^ (1) = 1.24, *p* = .26), highlighting that our blinding procedures were successful.

### The Effect of the Combined Intervention on Immediate Emotion Regulation

#### High-Frequency Heart Rate Variability Results

Active rTMS (EMM_*lg_HF-HRV*_ = 1.72, *S*.*E*. = .02) led to significantly higher HF-HRV when compared to sham (EMM_*lg_HF-HRV*_= 1.55, *S*.*E*. = .02; *F*[1, 19.00] = 69.38, *p* <.001, *d* = .43, *p*_FDR_ = .01), indicating enhanced emotion regulation. Emotion regulation as measured by HF-HRV was most effective at the beginning of the regulation period and declined by the last 120 s (*F*_*time*_[4, 24.92] = 4.04, *p* = 0.12, *p*_FDR_ = .03). Higher HF-HRV at baseline (*F*_*session_baseline*_[1, 26.20] = 135.27; *F*_pre-stimulus_baseline_[1, 30.59] = 483.84, *p*s <.001, *p*_FDR_ > .002) and smaller scalp-to-cortex distance (*F*[1, 18.85] = 17.04, *p*_FDR_ = .02) predicted higher HF-HRV during regulation (*p*s < .001). The time-by-condition interaction (*F*[4, 25.04] = 2.72, *p* = .053) showed significant differences between conditions at all time points (*p*s < .001; Figure 2), with the highest difference occurring between 120 s and 240 s (Δ_lg HF-HRV_ = .20; *d* = 0.50). The difference between active and sham was higher (Δ_lg_HF-HRV_ = .212; S.E. = .02; *d* = .53) in a secondary analysis excluding participants with high ERQ Reappraisal scores at baseline. Therefore, the enhancing effect of rTMS was stronger for those who didn’t habitually engage in reappraisal to manage negative emotions.

#### Self-Report Results

The experimental instructions (i.e., try to relax, remember a stressor, use CR) successfully modulated distress across conditions (*F*[2, 24.06] = 86.40, *p* < .001, *p*_FDR_ = .01). Follow up comparisons showed that participants significantly increased their distress after autobiographical emotion induction and significantly decreased it after the regulation period. There was no significant fixed effect for treatment condition (*F*[1,20.82] = 0.96, *p* = .34, *d* = .21, *p*_FDR_ = .04) or treatment condition-by-instruction interaction (*F*[2,24.00] = 0.04, *p* = .99). Baseline SUDS was a significant predictors of distress (*p*s < .001, *p*_FDR_ = .01). Scalp-to-cortex distance was not significantly related to SUDS after the FDR correction (*p* = 0.04, *p*_FDR_ = .036).When only including those with low ERQ Reappraisal, there was a trend for participants who received active versus sham rTMS to experience less subjective distress throughout the experiment (Δ_EMM_active-EMM_sham_ = −0.66; *F*[1, 11.47] = 3.85, *p* = .07, *d* = .37).

#### Regulation duration results

Twenty-six participants had at least one experimental condition in which their HR was above pre-stimulus baseline at the beginning of the regulation period. It took twice as long to return to HR baseline in the sham condition (M_sham_ =111.48 s, S.D. = 179.58) than in the active condition (M_active_ = 46.71 s, S.D. = 99.53), a difference that was significantly different (*F*_*treatment_condition*_[1, 48.16] = 4.85, *p* = 0.033, *p*_FDR_ = .034). The scalp-to-cortex distance was a significant predictor (*F*[1, 49.22] = 10.84, *p* < .002, *p*_FDR_ = .03), but baseline was not (*p* = .90). Larger scalp-to-cortex distance yielded significantly higher regulation duration (Parameter estimate = 0.06; SE = 0.02). The effect size of the difference between conditions was moderate (*d* = .48). Including only participants who self-reported low ERQ Reappraisal at baseline yielded a larger effect size (*d* = .63).

In summary, we found preliminary evidence that, when compared to CR+sham, active rTMS administered in conjunction with CR enhances physiological (HF-HRV) and behavioral indices of emotion regulation even after controlling for multiple comparisons. Smaller scalp-to-cortex distance was a significant predictor of improved emotion regulation (e.g., higher HF-HRV, lower regulation duration) consistent with our expectations. Effects of the active intervention were larger for those with low to moderate ERQ Reappraisal scores at baseline.

### The Near-Term Effect of the Combined Intervention on Psychopathology and Use of CR

#### Ambulatory Assessment Results

Of the 1512 calls placed, participants provided valid data for 1128 calls (74.6%). Participants indicated having used CR since the previous call in 537 instances (35.5% of answered calls) and reported SUDs above 0 in 739 calls (70.30% of answered calls). Participants who received active rTMS during their CR practice (EMM = .57, S.E. =.04) were more likely to use CR during the ambulatory week then participants who received sham stimulation (EMM = .45, S.E.=.04; Wald χ^*2*^ (1) = 5.72, *p* = .02, *p*_FDR_ = .03). Scalp-to-cortex distance (Parameter estimate = 0.11; SE = 0.03) and use of CR at intake (Parameter estimate = 0.42; SE = 0.10) were significant predictors of use of CR during the ambulatory week (Wald χ^*2*^_*distance*_ (1) = 10.63; Wald χ^*2*^_*ERQ_R*_ (1) = 19.16; *p*s < .001; *p*_FDR_ > .02).

Participants who received sham neurostimulation reported less distress during the ambulatory week (*F*_*condition*_[1, 302.96] = 11.65, *p* < .001, *p*_FDR_ = .02; Δ_SUDS_ = .48, S.E. = .13) when compared to participants who received active rTMS. There was a significant effect of baseline (*p* <0.001, *q*_FDR_ =0.003), and no significant effect of scalp-to-cortex distance. Excluding those with high use of CR at intake did not change the results.

#### Longitudinal Assessment Results

HLM models controlling for baseline and using a compound symmetry covariance structure found no significant differences between conditions for emotion dysregulation, reappraisal, overall psychopathology severity, and work and social functioning (*p*s > *p*_FDR_ > .04; see Suppl. Table 3 for means and SDs by assessment period). Baseline assessments predicted all outcomes (*p*s < .01; *q*_FDR_ > .02)

#### 1 Month Follow-Up Stressor Task Results

During the follow-up stressor task, participants were successful in modulating their subjective distress using a fourth autobiographical stressor and CR as a regulation technique (*F*_*time*_ [2, 40.87] *=* 15.30, *p* < .001, *q*_FDR_ = .01). Baseline SUDS predicted SUDS during the stressor task (*p* < .001, *q*_FDR_ = .01). There were no significant differences between condition in the success of regulation (*F*_*HF-HRV*_ [1, 18] *=* 0.08; *F*_*SUDS*_ [1, 40.52] *=* 0.18; *t*_*regulation_duration*_[12] = .89; *p*s > .05). When examining only those with low use of reappraisal at baseline, participants who had received active rTMS experienced marginally less self-reported distress during the stressor task when compared to participants receiving sham rTMS (*F*_*SUDS*_ [1, 21.82] *=* 4.10, *p* = 0.055, Δ_EMM_SUDS_ = -.95).

In summary, we found that active neurostimulation led to more use of CR the week after the intervention. Participants who received sham rTMS experienced less distress during the ambulatory assessment week. These results were significant even with FDR corrections.

## Discussion

We examined using a multi-method approach a brief, neuroscience-driven intervention for transdiagnostic emotional dysregulation. Specifically, we combined teaching and practice of a psychological skill, cognitive restructuring, with rTMS targeted towards the emotion regulation network using functional neuroimaging and neuronavigation. In addition, we examined the importance of skill proficiency at the beginning of the intervention on the emerging outcomes. As in our prior study(22), we found that active rTMS enhanced emotion regulation in the moment measured behaviorally and physiologically. When compared to sham, active rTMS over the left dlPFC in conjunction with CR practice increased HF-HRV and reduced regulation duration significantly, the difference corresponding to moderate effect sizes. The difference was due to the combined intervention: when we administered rTMS versus sham alone during the habituation portion, there were no differences between conditions. Therefore, this finding highlights that excitatory neurostimulation that targets the emotion regulation neural network enhances regulation of personalized stressors in transdiagnostic adults.

Our finding, expands existing evidence in the literature that neurostimulation can augment emotion regulation(20). Furthermore, it contributes to the literature that demonstrates performance enhancement effects of neurostimulation more broadly(24, 78-80). While the majority of performance optimization studies with neurostimulation targeted healthy adults, we provide emerging evidence that this augmentation effect is feasible to attain in dysregulated clinical samples and, therefore, that they may provide a promising avenue for treatment optimization.

Another important finding in our current study is the importance of functional targeting. In early intervention trials(81), neurostimulation was targeted by measuring 5 cm anterior to the spot identified as the rMT in the anterior-posterior plane. While this method is cost effective and easy to implement, more recent studies show that it may miss the dlPFC in over 30% of cases (82). A more precise method(83) that takes into account individual anatomy is the “beam” method which uses individual scalp landmarks such as F3 based on the International 10–20 System for placement of EEG recording electrodes(84). For most clear examinations of rTMS therapeutic effects, Luber et al. (2017) argue the importance of personalizing targeting with neuroimaging to ensure that the neuroplasticity induced by the rTMS hits the impaired circuit most likely to be responsible for symptoms. For example, targeting rTMS using a structural scan to identify the dlPFC versus the 5 cm rule may improve response rates in depression from 18% to 42% (although this difference was not statistically significant)(85). The superiority of functionally-guided rTMS over the beam methods has been demonstrated in a large meta-analysis on heathy controls(86), and in a theoretical examination of depression and the self-regulation circuitry(21). A similar case can be made for affect dysregulation.

In a prior study(22) we administered the same onetime combined CR and rTMS intervention to a transdiagnostic sample but we employed the “beam” method(84) to target rTMS. The intervention and procedures were no different, except that, in the current study, targeting was done using functional imaging. In the current study, we asked participants to undergo an autobiographical negative emotional induction in the scanner, and identified the sub-region within the left dlPFC with the highest *z* score when brain activity during cognitive emotion regulation was contrasted with emotional experience. In our prior study, targeting used scalp measurements to estimate where the dlPFC was located. The more sophisticated targeting in this study increased the effect size of the difference between active and sham for HF-HRV by 43% (from 0.30 in the first study to 0.43), and reduced the average regulation duration in the active condition by 58% (from 74.20 s to 46.71 s) (22). Although preliminary, this finding quantifies the potential gain of using functional targeting over the “beam” method.

We included in the study participants who reported different levels of proficiency with CR (defined using empirically based cutoffs). We were interested to examine whether outcomes varied depending on whether CR was a skills deficit or a strength. Although strengths-based approaches have a rich tradition in psychotherapy(87-89), most contemporary interventions assume that those who report emotion dysregulation lack the requisite skills to manage their emotions effectively(90-93). Nevertheless, even those with high emotion dysregulation have and can use adaptive regulation strategies under stress(1, 3, 94-96). Capitalizing on strengths may also lead to reduction in psychopathology more so that remediating deficits(97). In our study, we found that the greatest benefit came for participants who used CR very little before the intervention. All effect sizes were larger when excluding those with high or moderate to high use of CR at intake. Furthermore, when only examining participants with a skills deficit we found trends for significantly lower self-reported distress during the intervention and at the one month follow up for those who received active stimulation in conjunction with CR practice. Thus, our findings support the utility of a skills deficit model for transdiagnostic emotional dysregulation.

Scalp-to-cortex distance was a significant predictor of outcomes. Specifically, lower distance was associated with more desirable outcomes in the moment than higher distance. This finding highlights that the success of neurostimulation is sensitive to the individual brain anatomy and the resulting spatial distribution of the induced E-field(98). Others have also shown that the E-field strength is a key determinant of TMS-induced neural activation(99). Therefore, a next step in the optimization of this one-time intervention is the inclusion of E-field modeling in determining the strength of the magnetic field utilized for neurostimulation(100). An alternative approach would be the utilization of Stokes formula to account for scalp-to-cortex distance if structural imaging is available(68).

Our findings for near and long term effects of the combined intervention were mixed. Consistent with our previous study(22), we found that active neurostimulation over dlPFC increased use of CR during the week after the intervention when compared to sham. Contrary to our expectations and previous findings, participants in the sham condition reported less distress during the week. By the end of the week, no differences in self-report could be seen, and by one month after the intervention all participants were similar in their improvements in emotional dysregulation. Our lack of significant behavioral findings after a week may be due to the small sample size or to the small dose of active neurostimulation administered. Active stimulation may lead to more stress, although it’s likely as possible that participants randomized to the sham condition experienced more life stressors during the week following the intervention. A larger sample, control for number of stressors, and a higher dose of rTMS may clarify the effects of neurostimulation on long-term emotion regulation.

It is important to highlight that these results add to the growing literature aimed to understand and change emotional dysregulation transdiagnostically. In a prior treatment study (1), we recruited participants with affective disorders who self-reported high emotional dysregulation and conducted a behavioral intervention with this transdiagnostic sample (42% primarily depressed, 37% primary GAD, 15% primary social anxiety, 6% other anxiety). We found significant improvements in emotional regulation and psychopathology following an active versus a control behavioral treatment(1, 2). In both our prior(22) and current study combined, we enrolled 77 transdiagnostic participants with emotion dysregulation and found significant changes in several indices of regulation. A common set of neural dysfunctions connected to emotional dysregulation can also be seen across disorders(5). Thus, difficulties with emotion regulation cut across disorders and can be targeted by interventions independent of the diagnostic profile.

The study limitations include the small sample size and lack of matched control group for the skills training component. E-field modeling(101), which is recommended to optimize rTMS administration, was not employed. The rTMS protocol was borrowed from depression studies given the novel nature of the intervention. Nevertheless, the specific amount and spacing of the pulses administered may also be a limitation(102). Future studies are needed to replicate findings and refine procedures with larger samples.

In summary, our goal was to test a time-limited combined intervention for transdiagnostic emotional dysregulation. We found confirmatory evidence that rTMS augments behavioral skills training in the moment, and that the onetime session of combined rTMS-CR intervention targeted at a dlPFC component of the emotion regulation network generated effects that last as long as a week. Evidence of behavioral impact could not be found past a week which highlights the need for a larger study to replicate these findings. Emotional dysregulation is a key problem that cuts across psychiatric disorders and is associated with considerable burden of illness. Our findings provide important steps to address the critical need to develop new therapeutic approaches that integrate current knowledge about the neurophysiology of emotion regulation into clinical practice.

## Supporting information

consort checklist

## Data Availability

All data produced in the present study are available upon reasonable request to the authors

## Acknowledgements

Data from the present paper were also presented as part of several conference talks. This research and the completion of the manuscript were supported by a KL2 award granted to the first author by the National Center for Advancing Translational Sciences of the National Institutes of Health under Award Number 5KL2TR001115. The authors would like to thank the participants who took part in this study and to acknowledge Lisalynn Kelley, John Powers, PhD, M. Zachary Rosenthal, PhD, Kevin Haworth, PhD, Megan Renna, PhD, Simon Davis, PhD, Timothy Strauman, PhD, and our research assistants and the DUMC BIAC and BSRC staff for their contributions.

## Author Contributions

All authors contributed significantly to the present manuscript. Dr. Neacsiu conceptualized and conducted the study. Drs. LaBar, Appelbaum, and Smoski provided design input, helped problem solve study related issues throughout the study, and contributed to the manuscript preparation. Dr. Beynel conducted the neurostimulation procedures. Dr. Graner completed our fMRI preprocessing pipeline. Dr. Szabo served as the study doctor throughout the study.

## Data Availability Statement

All data is available upon request.

## References

1. Neacsiu AD, Eberle JW, Kramer R, Wiesmann T, Linehan MM. Dialectical behavior therapy skills for transdiagnostic emotion dysregulation: A pilot randomized controlled trial. Behav Res Ther. 2014;59:40–51.

2. Neacsiu AD, Rompogren J, Eberle JW, McMahon K. Changes in Problematic Anger, Shame, and Disgust in Anxious and Depressed Adults Undergoing Treatment for Emotion Dysregulation. Behav Ther. 2018;49(3):344–59.

3. Neacsiu AD, Rizvi SL, Linehan MM. Dialectical behavior therapy skills use as a mediator and outcome of treatment for borderline personality disorder. Behaviour Research and Therapy. 2010;48(9):832–9.

4. Neacsiu AD, Luber BM, Davis SW, Bernhardt E, Strauman TJ, Lisanby SH. On the Concurrent Use of Self-System Therapy and Functional Magnetic Resonance Imaging–Guided Transcranial Magnetic Stimulation as Treatment for Depression. The journal of ECT. 2018;34(4):266–73.

5. Zilverstand A, Parvaz MA, Goldstein RZ. Neuroimaging cognitive reappraisal in clinical populations to define neural targets for enhancing emotion regulation. A systematic review. NeuroImage. 2017;151:105–16.

6. Pico-Perez M, Radua J, Steward T, Menchon JM, Soriano-Mas C. Emotion regulation in mood and anxiety disorders: A meta-analysis of fMRI cognitive reappraisal studies. Prog Neuropsychopharmacol Biol Psychiatry. 2017;79(Pt B):96–104.

7. Buhle JT, Silvers JA, Wager TD, Lopez R, Onyemekwu C, Kober H, et al. Cognitive reappraisal of emotion: a meta-analysis of human neuroimaging studies. Cereb Cortex. 2014;24(11):2981–90.

8. Hölzel BK, Lazar SW, Gard T, Schuman-Olivier Z, Vago DR, Ott U. How does mindfulness meditation work? Proposing mechanisms of action from a conceptual and neural perspective. Perspectives on Psychological Science. 2011;6(6):537–59.

9. Fernandez KC, Jazaieri H, Gross JJ. Emotion Regulation: A Transdiagnostic Perspective on a New RDoC Domain. Cognit Ther Res. 2016;40(3):426–40.

10. Hawley LL, Padesky CA, Hollon SD, Mancuso E, Laposa JM, Brozina K, et al. Cognitive-behavioral therapy for depression using mind over mood: CBT skill use and differential symptom alleviation. Behavior Therapy. 2017;48(1):29–44.

11. Goldin PR, Ziv M, Jazaieri H, Werner K, Kraemer H, Heimberg RG, et al. Cognitive reappraisal self-efficacy mediates the effects of individual cognitive-behavioral therapy for social anxiety disorder. Journal of consulting and clinical psychology. 2012;80(6):1034.

12. Kaczkurkin AN, Foa EB. Cognitive-behavioral therapy for anxiety disorders: an update on the empirical evidence. Dialogues Clin Neurosci. 2015;17(3):337.

13. Powers JP, LaBar KS. Regulating emotion through distancing: A taxonomy, neurocognitive model, and supporting meta-analysis. Neuroscience and biobehavioral reviews. 2019;96:155–73.

14. Ochsner KN, Silvers JA, Buhle JT. Functional imaging studies of emotion regulation: a synthetic review and evolving model of the cognitive control of emotion. Annals of the New York Academy of Sciences. 2012;1251:E1–24.

15. Webb TL, Miles E, Sheeran P. Dealing with feeling: a meta-analysis of the effectiveness of strategies derived from the process model of emotion regulation. Psychological bulletin. 2012;138(4):775–808.

16. Denson TF, Grisham JR, Moulds ML. Cognitive reappraisal increases heart rate variability in response to an anger provocation. Motiv Emot. 2011;35(1):14–22.

17. Butler EA, Wilhelm FH, Gross JJ. Respiratory sinus arrhythmia, emotion, and emotion regulation during social interaction. Psychophysiology. 2006;43(6):612–22.

18. Di Simplicio M, Costoloni G, Western D, Hanson B, Taggart P, Harmer CJ. Decreased heart rate variability during emotion regulation in subjects at risk for psychopathology. Psychological Medicine. 2012;42(08):1775–83.

19. Mai S, Braun J, Probst V, Kammer T, Pollatos O. Changes in emotional processing following interoceptive network stimulation with rTMS. Neuroscience. 2019.

20. Feeser M, Prehn K, Kazzer P, Mungee A, Bajbouj M. Transcranial Direct Current Stimulation Enhances Cognitive Control During Emotion Regulation. Brain Stimulation. 2014;7(1):105–12.

21. Luber BM, Davis S, Bernhardt E, Neacsiu A, Kwapil L, Lisanby SH, et al. Using neuroimaging to individualize TMS treatment for depression: Toward a new paradigm for imaging-guided intervention. NeuroImage. 2017;148:1–7.

22. Neacsiu AD, Beynel L, Powers JP, Szabo ST, Appelbaum LG, Lisanby SH, et al. Enhancing Cognitive Restructuring with Concurrent Repetitive Transcranial Magnetic Stimulation for Transdiagnostic Psychopathology: A Proof of Concept Randomized Controlled Trial. medRxiv. 2021:2021.01.18.21250060.

23. Stefan K, Kunesch E, Cohen LG, Benecke R, Classen J. Induction of plasticity in the human motor cortex by paired associative stimulation. Brain. 2000;123(3):572–84.

24. Luber B, Stanford AD, Bulow P, Nguyen T, Rakitin BC, Habeck C, et al. Remediation of sleep-deprivation-induced working memory impairment with fMRI-guided transcranial magnetic stimulation. Cerebral cortex. 2008;18(9):2077–85.

25. Appelhans BM, Luecken LJ. Heart rate variability as an index of regulated emotional responding. Review of General Psychology. 2006;10(3):229–40.

26. Camm A, Malik M, Bigger J, Breithardt G, Cerutti S, Cohen R, et al. Heart rate variability: standards of measurement, physiological interpretation and clinical use. Task Force of the European Society of Cardiology and the North American Society of Pacing and Electrophysiology. Circulation. 1996;93(5):1043–65.

27. Geisler F, Kubiak T. Heart rate variability predicts selfUcontrol in goal pursuit. European Journal of Personality. 2009;23(8):623–33.

28. Ingjaldsson JT, Laberg JC, Thayer JF. Reduced heart rate variability in chronic alcohol abuse: relationship with negative mood, chronic thought suppression, and compulsive drinking. Biol Psychiatry. 2003;54(12):1427–36.

29. Demaree H, Schmeichel B, Robinson J, Everhart DE. Behavioural, affective, and physiological effects of negative and positive emotional exaggeration. Cognition & Emotion. 2004;18(8):1079–97.

30. Thayer JF, Åhs F, Fredrikson M, Sollers III JJ, Wager TD. A meta-analysis of heart rate variability and neuroimaging studies: implications for heart rate variability as a marker of stress and health. Neuroscience & Biobehavioral Reviews. 2012;36(2):747–56.

31. Fabes RA, Eisenberg N. Regulatory control and adults’ stress-related responses to daily life events. J Pers Soc Psychol. 1997;73(5):1107–17.

32. Fenton-O’Creevy M, Lins JT, Vohra S, Richards DW, Davies G, Schaaff K. Emotion regulation and trader expertise: Heart rate variability on the trading floor. Journal of Neuroscience, Psychology, and Economics. 2012;5(4):227.

33. Elliot AJ, Payen V, Brisswalter J, Cury F, Thayer JF. A subtle threat cue, heart rate variability, and cognitive performance. Psychophysiology. 2011;48(10):1340–5.

34. Neacsiu AD, Luber BM, Davis SW, Bernhardt E, Strauman TJ, Lisanby SH. On the Concurrent Use of Self-System Therapy and Functional Magnetic Resonance Imaging-Guided Transcranial Magnetic Stimulation as Treatment for Depression. J ECT. 2018;34(4):266–73.

35. Beynel L, Appelbaum LG, Luber B, Crowell CA, Hilbig SA, Lim W, et al. Effects of online repetitive transcranial magnetic stimulation (rTMS) on cognitive processing: A meta-analysis and recommendations for future studies. Neuroscience & Biobehavioral Reviews. 2019.

36. Sack AT, Cohen Kadosh R, Schuhmann T, Moerel M, Walsh V, Goebel R. Optimizing functional accuracy of TMS in cognitive studies: a comparison of methods. J Cogn Neurosci. 2009;21(2):207–21.

37. Faul F, Erdfelder E, Buchner A, Lang AG. Statistical power analyses using G*Power 3.1: tests for correlation and regression analyses. Behavior research methods. 2009;41(4):1149–60.

38. First MB, Williams JB, Karg RL. Structured Clinical Interview for DSM-5 Disorders (Research Version: SCID-5-RV). Washington, DC: American Psychiatric Publishing; 2015.

39. First MB, Williams JB, Benjamin LS, Spitzer RL. SCID-5-PD: Structured clinical interview for DSM-5® personality disorders: American Psychiatric Association Publishing; 2016.

40. Gratz KL, Roemer L. Multidimensional Assessment of Emotion Regulation and Dysregulation: Development, Factor Structure, and Initial Validation of the Difficulties in Emotion Regulation Scale. J Psychopathol Behav Assess. 2004;26(1):41–54.

41. Gross JJ, John OP. Individual differences in two emotion regulation processes: Implications for affect, relationships, and well-being. Journal of Personality and Social Psychology. 2003;85(2):348–62.

42. Dunn LM. PPVT-revised manual. Circle Pines, MN: American Guidance Service; 1981 1981.

43. Mundt JC, Marks IM, Shear MK, Greist JM. The Work and Social Adjustment Scale: a simple measure of impairment in functioning. The British Journal of Psychiatry. 2002;180(5):461–4.

44. Lambert MJ, Burlingame GM, Umphress V, Hansen NB, Vermeersch DA, Clouse GC, et al. The reliability and validity of the Outcome Questionnaire. Clinical Psychology & Psychotherapy: An International Journal of Theory and Practice. 1996;3(4):249–58.

45. Pitman RK, Orr SP, Forgue DF, de Jong JB, Claiborn JM. PSychophysiologic assessment of posttraumatic stress disorder imagery in vietnam combat veterans. Arch Gen Psychiatry. 1987;44(11):970–5.

46. Schmahl CG, Elzinga BM, Ebner UW, Simms T, Sanislow C, Vermetten E, et al. Psychophysiological reactivity to traumatic and abandonment scripts in borderline personality and posttraumatic stress disorders: a preliminary report. Psychiatry Res. 2004;126(1):33–42.

47. Kross E, Davidson M, Weber J, Ochsner K. Coping with emotions past: the neural bases of regulating affect associated with negative autobiographical memories. Biol Psychiatry. 2009;65(5):361–6.

48. Talarico JM, LaBar KS, Rubin DC. Emotional intensity predicts autobiographical memory experience. Mem Cognit. 2004;32(7):1118–32.

49. Greenberg DL, Rice HJ, Cooper JJ, Cabeza R, Rubin DC, LaBar KS. Co-activation of the amygdala, hippocampus and inferior frontal gyrus during autobiographical memory retrieval. Neuropsychologia. 2005;43(5):659–74.

50. Powers JP, Graner JL, LaBar KS. Multivariate Patterns of Posterior Cortical Activity Differentiate Forms of Emotional Distancing. Cerebral cortex. 2019.

51. Scott NW, McPherson GC, Ramsay CR, Campbell MK. The method of minimization for allocation to clinical trials. a review. Control Clin Trials. 2002;23(6):662–74.

52. Taves DR. Minimization: a new method of assigning patients to treatment and control groups. Clin Pharmacol Ther. 1974;15(5):443–53.

53. Esteban O, Markiewicz CJ, Blair RW, Moodie CA, Isik AI, Erramuzpe A, et al. fMRIPrep: a robust preprocessing pipeline for functional MRI. Nature methods. 2019;16(1):111.

54. Smith SM. Fast robust automated brain extraction. Hum Brain Mapp. 2002;17(3):143–55.

55. Jenkinson M, Beckmann CF, Behrens TE, Woolrich MW, Smith SM. Fsl. NeuroImage. 2012;62(2):782–90.

56. Avants BB, Tustison NJ, Song G, Cook PA, Klein A, Gee JC. A reproducible evaluation of ANTs similarity metric performance in brain image registration. NeuroImage. 2011;54(3):2033–44.

57. Linehan M. DBT? Skills training manual: Guilford Publications; 2014.

58. Beck A, Emery G, Greenberg R. Anxiety Disorders and Phobias: A Cognitive Perspective Basic Books. New York. 1985.

59. Shurick AA, Hamilton JR, Harris LT, Roy AK, Gross JJ, Phelps EA. Durable effects of cognitive restructuring on conditioned fear. Emotion. 2012;12(6):1393–7.

60. Beck AT, Dozois DJ. Cognitive therapy: current status and future directions. Annu Rev Med. 2011;62:397–409.

61. Ochsner KN, Bunge SA, Gross JJ, Gabrieli JDE. Rethinking feelings: an FMRI study of the cognitive regulation of emotion. J Cogn Neurosci. 2002;14(8):1215–29.

62. Kendall PC. Coping cat workbook: Workbook Pub; 2006.

63. Rossi S, Hallett M, Rossini PM, Pascual-Leone A, Group SoTC. Safety, ethical considerations, and application guidelines for the use of transcranial magnetic stimulation in clinical practice and research. Clin Neurophysiol. 2009;120(12):2008–39.

64. Wassermann EM. Risk and safety of repetitive transcranial magnetic stimulation: report and suggested guidelines from the International Workshop on the Safety of Repetitive Transcranial Magnetic Stimulation, June 5–7, 1996. Electroencephalography and Clinical Neurophysiology/Evoked Potentials Section. 1998;108(1):1–16.

65. Smith JE, Peterchev AV. Electric field measurement of two commercial active/sham coils for transcranial magnetic stimulation. Journal of neural engineering. 2018;15(5):054001.

66. Stiglmayr C, Schmahl C, Bremner JD, Bohus M, Ebner-Priemer U. Development and psychometric characteristics of the DSS-4 as a short instrument to assess dissociative experience during neuropsychological experiments. Psychopathology. 2009;42(6):370–4.

67. Lee E, Duffy W, Hadimani R, Waris M, Siddiqui W, Islam F, et al. Investigational effect of brain-scalp distance on the efficacy of transcranial magnetic stimulation treatment in depression. IEEE Transactions on Magnetics. 2016;52(7):1–4.

68. Stokes MG, Chambers CD, Gould IC, Henderson TR, Janko NE, Allen NB, et al. Simple metric for scaling motor threshold based on scalp-cortex distance: application to studies using transcranial magnetic stimulation. J Neurophysiol. 2005;94(6):4520–7.

69. Powers JP, Davis SW, Neacsiu AD, Beynel L, Appelbaum LG, LaBar KS. Examining the Role of Lateral Parietal Cortex in Emotional Distancing Using TMS. Cognitive, Affective, & Behavioral Neuroscience. 2020;20(5):1090–102.

70. Molenberghs G, Verbeke G. Linear mixed models for longitudinal data: Springer; 2000.

71. Gibbons RD, Hedeker D, DuToit S. Advances in analysis of longitudinal data. Annu Rev Clin Psychol. 2010;6:79–107.

72. Benjamini Y, Hochberg Y. Controlling the false discovery rate: a practical and powerful approach to multiple testing. Journal of the Royal statistical society: series B (Methodological). 1995;57(1):289–300.

73. Feingold A. Effect sizes for growth-modeling analysis for controlled clinical trials in the same metric as for classical analysis. Psychol Methods. 2009;14(1):43.

74. Cohen J. Statistical power analysis for the behavioral sciences Rev. ed. ed. New York, NY: Academic Press; 1977.

75. Raudenbush SW, Bryk AS. Hierarchical linear models: Applications and data analysis methods: Sage; 2002.

76. Zeger SL, Liang K-Y. Longitudinal data analysis for discrete and continuous outcomes. Biometrics. 1986:121–30.

77. Zeger SL, Liang K-Y, Albert PS. Models for longitudinal data: a generalized estimating equation approach. Biometrics. 1988:1049–60.

78. Luber B, Steffener J, Tucker A, Habeck C, Peterchev AV, Deng ZD, et al. Extended remediation of sleep deprived-induced working memory deficits using fMRI-guided transcranial magnetic stimulation. Sleep. 2013;36(6):857–71.

79. Curtin A, Ayaz H, Tang Y, Sun J, Wang J, Tong S. Enhancing neural efficiency of cognitive processing speed via training and neurostimulation: An fNIRS and TMS study. NeuroImage. 2019;198:73–82.

80. Grimaldi D, Papalambros NA, Zee PC, Malkani RG. Neurostimulation techniques to enhance sleep and improve cognition in aging. Neurobiol Dis. 2020;141:104865.

81. George MS, Wassermann EM, Williams WA, Callahan A, Ketter TA, Basser P, et al. Daily repetitive transcranial magnetic stimulation (rTMS) improves mood in depression. Neuroreport: An International Journal for the Rapid Communication of Research in Neuroscience. 1995.

82. Johnson KA, Baig M, Ramsey D, Lisanby SH, Avery D, McDonald WM, et al. Prefrontal rTMS for treating depression: location and intensity results from the OPT-TMS multi-site clinical trial. Brain Stimulation. 2013;6(2):108–17.

83. Mir-Moghtadaei A, Caballero R, Fried P, Fox MD, Lee K, Giacobbe P, et al. Concordance between BeamF3 and MRI-neuronavigated target sites for repetitive transcranial magnetic stimulation of the left dorsolateral prefrontal cortex. Brain Stimulation. 2015;8(5):965–73.

84. Beam W, Borckardt JJ, Reeves ST, George MS. An efficient and accurate new method for locating the F3 position for prefrontal TMS applications. Brain Stimulation. 2009;2(1):50–4.

85. Fitzgerald PB, Hoy K, McQueen S, Maller JJ, Herring S, Segrave R, et al. A randomized trial of rTMS targeted with MRI based neuro-navigation in treatment-resistant depression. Neuropsychopharmacology. 2009;34(5):1255–62.

86. Beynel L, Campbell E, Naclerio M, Galla JT, Ghosal A, Michael AM, et al. The effect of functionally-guided-connectivity-based rTMS on amygdala activation. bioRxiv. 2020:2020.10.13.338483.

87. Goldiamond I. Toward a constructional approach to social problems: ethical and constitutional issues raised by applied behavior analysis. Behaviorism. 1973;2(1):1–84.

88. Weick A, Rapp C, Sullivan WP, Kisthardt W. A strengths perspective for social work practice. Social Work. 1989;34:350–4.

89. De Shazer S. Keys to solution in brief therapy New York: Ww Norton; 1985.

90. Mennin DS. Emotion Regulation Therapy: An Integrative Approach to Treatment-Resistant Anxiety Disorders. J Contemp Psychother. 2006;36(2):95–105.

91. Barlow DH, Farchione TJ, Fairholme CP, Ellard KK, Boisseau CL, Allen LB, et al. Unified protocol for transdiagnostic treatment of emotional disorders: Therapist guide: Oxford University Press, USA; 2010.

92. Linehan M. Cognitive-behavioral Treatment of Borderline Personality Disorder: Guilford Press; 1993 1993. 584 p.

93. Gratz KL, Gunderson JG. Preliminary Data on an Acceptance-Based Emotion Regulation Group Intervention for Deliberate Self-Harm Among Women With Borderline Personality Disorder. Behavior Therapy. 2006;37(1):25–35.

94. Gruber J, Harvey AG, Gross JJ. When trying is not enough: emotion regulation and the effort-success gap in bipolar disorder. Emotion. 2012;12(5):997–1003.

95. Gruber J, Kogan A, Mennin D, Murray G. Real-world emotion? An experience-sampling approach to emotion experience and regulation in bipolar I disorder. Journal of abnormal psychology. 2013;122(4):971–83.

96. Neacsiu AD, Smith M, Fang CM. Challenging assumptions from emotion dysregulation psychological treatments. J Affect Disord. 2017;219:72–9.

97. Cheavens JS, Strunk DR, Lazarus SA, Goldstein LA. The compensation and capitalization models: a test of two approaches to individualizing the treatment of depression. Behav Res Ther. 2012;50(11):699–706.

98. Peterchev AV, Wagner TA, Miranda PC, Nitsche MA, Paulus W, Lisanby SH, et al. Fundamentals of transcranial electric and magnetic stimulation dose: Definition, selection, and reporting practices. Brain Stimulation. 2012;5(4):435–53.

99. Aberra AS, Wang B, Grill WM, Peterchev AV. Simulation of transcranial magnetic stimulation in head model with morphologically-realistic cortical neurons. Brain Stimul. 2020;13(1):175–89.

100. Janssen AM, Oostendorp TF, Stegeman DF. The coil orientation dependency of the electric field induced by TMS for M1 and other brain areas. J Neuroeng Rehabil. 2015;12.

101. Deng Z-D, Lisanby SH, Peterchev AV. Electric field depth–focality tradeoff in transcranial magnetic stimulation: simulation comparison of 50 coil designs. Brain Stimulation. 2013;6(1):1–13.

102. Cash RF, Dar A, Hui J, De Ruiter L, Baarbé J, Fettes P, et al. Influence of inter-train interval on the plastic effects of rTMS. Brain Stimulation. 2017;10(3):630–6.

