## Supplemental Table 3 for "Enhancing Cognitive Restructuring with Concurrent fMRI-guided Neurostimulation for Emotional Dysregulation: A Randomized Controlled Trial"

Table 2. Means and SDs for longitudinal outcomes for all participants and broken by condition

|  |  | | OQ-45 | DERS | ERQ-R | WSAS |
| --- | --- | --- | --- | --- | --- | --- |
| All | Intake (N = 27) | | 71.16 (25.44) | 109.96 (17.43) | 4.26 (1.33) | 19.31 (7.77) |
|  | 1 week FU (N = 26) | | 73.58 (26.15) | 99.08 (24.92) | 4.70 (1.29) | 14.89 (9.94) |
|  | 1 month FU (N = 23) | | 65.44 (33.12) | 86.78 (23.10) | 4.92 (1.43) | 9.22 (9.43) |
| CR + active rTMS | Intake (N = 14) | | 74.69 (24.86) | 109.57 (17.02) | 4.10 (1.35) | 21.62 (6.92) |
|  | 1 week FU (N = 14) | | 78.79 (24.81) | 107.43 (24.69) | 4.73 (1.45) | 17.29 (9.47) |
|  | 1 month FU (N = 12) | | 65.08 (30.34) | 87.83 (18.98) | 4.81 (1.71) | 10.17 (8.05) |
| CR + sham rTMS | Intake (N = 13) | | 67.33 (26.60) | 110.38 (18.54) | 4.44 (1.33) | 17.00 (8.14) |
|  | 1 week FU (N = 12) | | 67.50 (27.42) | 89.33 (22.32) | 4.67 (1.14) | 12.08 (10.12) |
|  | 1 month FU (N = 11) | | 65.82 (37.43) | 85.64 (27.83) | 5.05 (1.13) | 8.18 (11.04) |
| Normative  data | | > 63 = clinical impairment; reliable change = 17 points | | < 80 = individuals not in treatment | > 4.67 – non-clinical | 10-20 clinical impairment  >20 serious psychopathology |

*Note.* FU = follow up; CR= Cognitive restructuring training and practice on autobiographical stressors. SD = standard deviation; OQ-45 = Outcome Questionnaire-45 total score; DERS = Difficulties in Emotion Regulation Scale Total Score; ERQ-R = Emotion Regulation Questionnaire- Reappraisal Scale; WSAS = Work and Social Adjustment Scale
