## supplemental table 1 for "Enhancing Cognitive Restructuring with Concurrent fMRI-guided Neurostimulation for Emotional Dysregulation: A Randomized Controlled Trial"

| Supplementary Table 1. *Breakdown between conditions by CR Proficiency skill in the ITT sample* | | | | |
| --- | --- | --- | --- | --- |
| Proficiency with cognitive restructuring |  | All  (*n* = 31) | Active Left  (*n* = 16) | Sham  (*n* = 15) |
| LOW | ERQ Reappraisal | 3.47 (0.89) | 3.37 (0.92) | 3.57 (0.88) |
|  | DERS Total | 110.90 (18.34) | 111.20 (19.01) | 110.56 (18.70) |
|  | N | 19 | 10 | 9 |
| MODERATE | ERQ Reappraisal | 5.13 (0.24) | 5.13 (0.22) | 5.11 (0.35) |
|  | DERS Total | 107.00 (8.91) | 106.40 (7.73) | 108.00 (12.49) |
|  | N | 8 | 5 | 3 |
| HIGH | ERQ Reappraisal | 6.13 (0.29) | 6.50 | 6.00 (0.16) |
|  | DERS Total | 108.25 (22.78) | 90.00 | 114.33 (23.59) |
|  | N | 4 | 1 | 3 |

*Note.* ERQ = Emotion Regulation Questionnaire; DERS = Difficulties in emotion regulation scale
