## supplemental table 2 for "Enhancing Cognitive Restructuring with Concurrent fMRI-guided Neurostimulation for Emotional Dysregulation: A Randomized Controlled Trial"

Table 1. *Demographics and Clinical Descriptives by Group*

|  | Active (*n* = 16) | Sham (*n* = 13) | Unrandomized (*n* = 2) |
| --- | --- | --- | --- |
| Mean age (SD) | 32.87 (13.99) | 35.77 (13.50) | 28.00 (14.14) |
| Female gender (%) | 81.25 | 84.62 | 50.00 |
| Latinx Background (%) | 6.25 | 15.38 | 0.00 |
| Racial Background (%) |  |  |  |
| Asian/Asian American | 18.75 | 7.69 | 0.00 |
| Black/African American | 18.75 | 7.69 | 0.00 |
| Native American, American Indian, or Alaskan Native | 6.25 | 0.00 | 0.00 |
| White/Caucasian | 56.25 | 69.23 | 100.00 |
| Multiracial | 0.00 | 15.38 | 0.00 |
| Relationship Status (%) |  |  |  |
| Single, never married | 68.75 | 53.85 | 50.00 |
| Married | 12.50 | 23.08 | 50.00 |
| Divorced | 6.25 | 0.00 | 0.00 |
| Living with Partner | 12.50 | 23.08 | 0.00 |
| Educational Attainment (%) |  |  |  |
| GED | 6.25 | 0.00 | 0.00 |
| High School Graduate | 12.50 | 0.00 | 50.00 |
| Some College | 12.50 | 30.77 | 0.00 |
| College Graduate | 43.75 | 53.85 | 0.00 |
| Some Graduate School | 6.25 | 7.69 | 0.00 |
| Master’s Degree | 12.50 | 0.00 | 50.00 |
| Doctoral Degree | 6.25 | 0.00 | 0.00 |
| Post-High School Business/Technical Training | 0.00 | 7.69 | 0.00 |
| Household Income (%) |  |  |  |
| $0-$10,000 | 12.50 | 15.38 | 0.00 |
| $10,001-$20,000 | 12.50 | 15.38 | 0.00 |
| $20,001-$40,000 | 25.00 | 23.08 | 50.00 |
| $40,001 - $65,000 | 18.75 | 23.08 | 0.00 |
| $65,001-$100,000 | 6.25 | 7.69 | 0.00 |
| More than $100,000 | 25.00 | 15.38 | 50.00 |
| Total # of Diagnoses, Current (SD) | 4.94 (2.08) | 5.15 (2.30) | 3.50 (0.71) |
| Total # of Diagnoses, Lifetime (SD) | 3.13 (1.75) | 3.46 (1.76) | 3.50 (0.71) |
| Current Disorders (%) |  |  |  |
| Anxiety Disorders | 87.50 | 92.31 | 100.00 |
| Bipolar Disorders | 50.00 (n = 2) | 0.00 (n = 2) | No data |
| Depressive Disorders | 28.57 (n = 14) | 50.00 (n = 10) | No data |
| Lifetime Disorders (%) |  |  |  |
| Anxiety Disorders | 87.50 | 92.31 | 100.00 |
| Depressive Disorders | 93.75 | 84.62 | 0.00 |
| Substance Use Disorders | 25.00 | 7.69 | 0.00 |
| Antisocial PD (%) | 0.00 (n = 14) | 0.00 | 0.00 (n = 1) |
| Avoidant PD (%) | 14.29 (n = 14) | 15.38 | 0.00 (n = 1) |
| Borderline PD (%) | 21.43 (n = 14) | 7.69 | 100.00 (n = 1) |
| Histrionic PD (%) | 0.00 (n = 14) | 0.00 | 0.00 (n = 1) |
| Obsessive-Compulsive PD (%) | 7.14 (n = 14) | 7.69 | 0.00 (n = 1) |
| Any PD (%) |  |  |  |

*Note:* PD = Personality Disorder
