## supplementary methods for "Enhancing Cognitive Restructuring with Concurrent fMRI-guided Neurostimulation for Emotional Dysregulation: A Randomized Controlled Trial"

***Supplemental Information***

### Supplemental Methods and Materials

### Participants and Procedures

Interested participants completed an online screen and were subsequently called for additional screening if they did not report meeting any exclusion criteria. Participants met criteria for at least one of the following diagnoses according to the Structured Clinical Interview (SCID-5) for *DSM-5* disorders (1): major depressive disorder (MDD; *n* = 5), persistent depressive disorder (PDD; *n* = 7), premenstrual dysphoric disorder (PMDD; *n =* 5), bipolar II disorder (*n* = 1), other depressive disorder (*n* = 1), panic disorder (*n* = 1), agoraphobia (*n* = 2), social anxiety disorder (SAD; *n* = 13), generalized anxiety disorder (GAD; *n* = 20), specific phobia (SP; *n =* 8), other specified anxiety disorder (OASD; *n* = 1), obsessive compulsive disorder (OCD; *n* = 2), body dysmorphic disorder (BDD; *n =* 5), hoarding disorder (*n* = 1), trichotillomania (*n* = 1), excoriation disorder (*n* = 3), bulimia nervosa (BN; *n* = 2), binge eating (*n* = 2), other eating disorder (*n* = 1), somatic symptom disorder (*n* = 1), adult attention deficit hyperactivity disorder (ADHD; *n* = 3), intermittent explosive disorder (IEE; *n* = 1), acute stress disorder (*n* = 1), posttraumatic stress disorder (PTSD; *n* = 7), other stress disorder (*n* = 4), adjustment disorder (*n* = 1).

High emotional dysregulation was operationally defined as a total score of 89 or higher on the Difficulties in Emotion Regulation Scale (DERS)(2). The cutoff was computed by pooling DERS total scores from 355 screens we completed in a previous study where we advertised for adults with high emotional dysregulation (3) and from samples in published studies before summer 2017 (2, 4-14). The pooled mean for the DERS total score (range: 36-170) across 3359 adults in 13 studies was 89.45 (*SD_pooled_* = 22.23). Therefore, we defined high emotion dysregulation as being having a total DERS score higher than 89. Intent to treat participants scored on average 109.55 (SD = 16.59) on the DERS, with no difference between conditions, *t*(29) = -0.40, *p* = .69.

The sample was stratified using the Emotion Regulation Questionnaire (ERQ)(15) cognitive reappraisal subscale (range: 1-7). To establish cutoffs for low, moderate and high reappraisal proficiency, we pooled the means and standard deviations from 18 studies (N = 4331 participants) published before 2014 that reported ERQ Reappraisal scores in adult samples (15-32). We considered adults who scored below this pooled mean (*M_pooled_* = 4.70; *SD_pooled_* = 0.99) to have “low use” of CR strategies. Adults with ERQ reappraisal scores between 4.7 and 5.7 (one SD_pooled_ over pooled mean) were categorized as having moderate proficiency with CR, and adults who scored higher than 5.7 on ERQ reappraisal were considered to have high proficiency with CR. ITT participants scored an average of 4.24 (SD = 1.25) on the ERQ cognitive reappraisal subscale, with no significant difference in scores between treatment conditions (*t*[29] = -0.56, *p* = .58). There break down in CR proficiency between groups is presented in Supplementary Table 1.

Participants were excluded if they were younger than 18 and older than 65; scored below the DERS cutoff; did not report any mental health difficulties; endorsed current substance or alcohol abuse or a history of psychosis; were currently in (or planning to start) CBT; needed immediate hospitalization; were at high risk for suicide; were at increased risk for seizure during neuromodulation because of medication, neurological disorder, brain injury, or family history; could not do MRI (weight, claustrophobia, metal in the body). Pregnant women, homeless individuals, individuals who could not travel to Duke, and who did not understand English or had low verbal IQ (33) and could not understand the intervention were also excluded. Participants unwilling to answer the ambulatory calls were also excluded. Two participants were excluded because they could not comply with the MRI instructions and therefore did not have usable data for the study. One of these participants was excluded after randomization, and their randomization was replaced to preserve balance (See CONSoRT Figure 1 for additional details.)

#### Intake Session

Qualified participants worked with the assessor to identify four autobiographical stressors to be used for emotional induction during intervention and follow up. Using the method developed by Pitman and colleagues (34) and used in studies of adults with affective disorders (35), we created four negative emotional arousal scripts, three of which were used in a random order during the intervention day and one during the 1-month follow-up. In brief, the assessor asked the participant to describe three moderate stressors from the past month, as well as a stressor that tended to reoccur for them. Participants wrote a description of each event, and the assessor worked with the participant to establish a clear story for each event that could be recited in ~40 seconds. Following the assessment, these scripts were narrated by the PI and digitally recorded into .wav audio files to be presented during the following sessions. Participants identified 10 negative and four neutral lifetime memories for autobiographical emotional induction during the fMRI session.

Autobiographical memories as means of emotional induction during MRI, intervention and follow up were chosen to more closely mirror the clinical processes that are targeted with therapeutic interventions.

#### Imaging sessions

The MRI sessions lasted 120 minutes and happened approximately one week before and after the intervention session. During the first hour, participants could go through a mock MRI session to familiarize themselves with the scanning environment in a deactivated scanner and ask any questions. They were also tested for pregnancy and participated in a training session for the strategies to use during the scan. Following standardized training, participants practiced the feel, distance, and reframe strategies on two situations by saying out loud what they would think in order for the experimenter to check comprehension. Strategies were covered sufficiently for the participant to be able to engage in the targeted behavior during the scan, but not in depth such that the participant learned new ways to downregulate emotions. The in-depth training of distancing and reframing was kept for the intervention session. Next, participants moved into the MRI scanner (**Figure 2B**). During the four runs of functional acquisitions, participants performed 40 trials of the emotion regulation task. Each trial of the task consisted of sequential presentations of: (1) a jittered cross (passive baseline), an arrow task (active baseline), a memory cue word (negative/neutral), a strategy word (“Feel”, “Distance”, or “Reframe”), a rating of distress. The arrow task consisted of three arrows appearing on the screen, one after another, for 3 seconds each. Each arrow pointed either to the left or to the right. Participants were asked to respond to the direction of each arrow via button press (index finger button for left, middle finger button for right). Following the arrow task, one of the memory cue words (in random order) was presented on the screen for 5 seconds. During training, participants had been instructed to use this time to bring the cued memory to mind. Following each memory cue word, a strategy word (one of “Feel”, “Distance”, or “Reframe”) was presented for 10 seconds. During these 10 seconds, participants were expected to either fully experience emotions associated with the cued memory (‘Feel’) or to try to downregulate them using either distancing or reframing CR strategies (“Distance”, “Reframe”, respectively). After these 10 seconds, participants were presented with a rating scales on which to report their current subjective units of distress (0-9; 10 s).

Visual stimuli were back projected onto a screen located at the foot of the MRI bed using an LCD projector. Participants viewed the screen via a mirror system located on the head coil and the start of each run was electronically synchronized with the MRI acquisition computer. Behavioral responses were recorded with a 4-key fiber-optic response box (Resonance Technology, Inc.). Scanner noise was reduced with ear plugs, and head motion was minimized with foam pads placed on both sides of the participants’ head. When necessary, vision was corrected using MRI-compatible lenses that matched the distance prescription used by the participant.

***Randomization***

Participants were matched according to sex, use of psychotropic medication, and proficiency with reappraisal using the QMinim software (<http://rct.mui.ac.ir/q/index.php>) by an independent staff member and kept blind from the assessor, PI and participant.

***Neurostimulation target identificatio****n:* ***additional information on preprocessing steps***

Structural and functional data were first examined with MRIQC and preprocessed with fMRIprep1.1.1 (36).Preprocessing of the high-resolution, T1-weighted anatomical images included intensity correction using N4BiasFieldCorrection (37), skull-stripping, and spatial normalization to the ICBM 152 Nonlinear Asymmetrical template v2009c (38). Initial preprocessing of the functional image data included slice time correction, calculation of motion correction transforms, and calculation of the transform for spatial registration to the high-resolution T1-weighted image. The fMRI data were then moved to standard space by applying the concatenated motion-correction transforms, fMRI-to-T1 transform, and T1-to-ICBM transform. The non-aggressive variant of ICA-based automatic removal of motion artifacts (AROMA) (39) was also applied to the fMRI data. After this preprocessing by fMRIprep, the fMRI data were skull-stripped (FSL’s Brain Extraction Tool) (40)and underwent high-pass temporal filtering with a 100-second cutoff (FSL’s fslmaths).

At the first level, functional data were analyzed as individual runs, using FSL’s FEAT. Model regressors were created for the following: presentation of the attentional arrow task (duration: 9s), presentation of the negative and neutral memory cues (5s), presentation of the “distance” strategy word (10s), presentation of the “reframe” strategy word (10s), presentation of the “feel” strategy word after the negative memory cues (“Feel Negative”; 10s), presentation of the “feel” strategy word after the neutral memory cures (“Feel Neutral”; 10s), presentation of the ratings of emotional state (10s). The onset and duration of each event was recorded by the Matlab script used to present the functional task and these were formatted for use in FSL.

#### Intervention Session: additional details on intervention training

During the CR training, participants were told that thoughts and emotions are interconnected and one validated way to change emotional experiences is to think less toxic thoughts in situations that prompt the emotions (41-44). Once one is experiencing a negative emotion, that they want to change, and they are willing to put effort towards changing it, one can choose between distancing and reframing tactics to reduce the intensity and the duration of an emotion. In teaching *distancing*, adopting a detached and unemotional attitude was introduced as objective distancing (45). Distancing by using time and space were also discussed and practiced on standardized examples (46).

Second, participants were taught *reframing* using an adapted version of existing paradigms (47-49). Specifically, we emphasized with pictures and examples the relationship between thoughts and emotions in ambiguous situations, identified effectiveness as the objective in an emotional situation, and instructed participants to find interpretations that are less toxic in order to be effective rather than right when upset. Participants learned to think about elements of the situation that they did not pay attention to or information that was missing, and to reframe their cognitions based on the full picture with an eye towards effectiveness. Participants were also taught to examine the worst-case scenario, the probability of it occurring, and the likelihood of survival if it occurred. The CR training ended with a quiz that included definitions as well as fake scenarios to practice on.

#### Intervention Session: additional details on concurrent rTMS and CR practice

The TMS-CR intervention started with a 600 s habituation period during which active or sham rTMS was applied while participants were listening to white noise via headphones. Participants were not instructed to think of anything in particular during this time. Then, the intervention proceeded as follows: (1) participants sat quietly for a 300 s pre-stimulus baseline while listening to white noise, (2) participants were instructed to imagine as vividly as possible one of the stressful experiences constructed at intake, (3) the stressor was heard via headphones followed by silence when participants were instructed to continue to imagine the stressful situation (120 s total), and (4) instructions in reducing distress using CR followed. The rTMS began within 10 seconds after the CR instructions appeared. Reminders of reframing and distancing appeared in random order 180 s and 360 s post-onset of stimulation. White noise was played throughout when instructions were not presented. After 600 s, there was a break, followed by a second and third administration of the stressor task using the procedures outlined above, but with a different personalized stressor recording each time. The order of the stressors that the participants described during the clinical assessment was randomized. After each baseline, stress induction, and regulation period the participant was asked to rate subjective units of distress (SUDS). At the end of the intervention, manipulation check questions ensuring compliance with the instructions and asking participants to guess the blind were administered.

**Measures**

***Diagnostic Assessment***

The SCID-5 (1) and SCID-PD (50) have demonstrated high diagnostic accuracy (83%) and strong inter-rater reliability (.85 during training and .76 at a Quality Assurance check) (51). Participants were led through both structured interviews by either the first author (65.5% of cases) or one of four trained diagnostic assessors under the supervision of the first author. In the cases where the first author did not conduct the interview, she reviewed in detail with the assessor the questions asked to confirm diagnostic profile. In case of disagreement, she reassessed the disorder at the next visit.

***Self-reports***

**Emotion Regulation Questionnaire (ERQ).** The ERQ (15) is a 10‐item scale that assesses two cognitive emotion regulation strategies: expressive suppression and cognitive reappraisal. The items use a 7‐point Likert scale to identify how likely one is to use each strategy to alter positive and negative emotions with responses ranging from one (*strongly disagree*) to 7 (*strongly agree*). The ERQ has good psychometric properties, including high internal reliability and validity (15, 18). In this study, Cronbach alpha at intake for ERQ Reappraisal was .92. The correlation between phone screen and intake reappraisal scores for 86 participants was .58. ERQ suppression was collected, but not examined.

**Difficulties in Emotion Regulation Scale (DERS).** The DERS (2) is a 36-item instrument that assesses typical levels of emotion dysregulation across six domains (awareness, nonacceptance, strategies, goal oriented behaviors, impulsivity, and emotional clarity). Participants respond on a Likert scale ranging from 1 (*almost never*) to 5 (*almost always*). In the original measure development paper, the DERS was found to have high internal consistency (α = .93), good test-retest reliability (*r* = .88, *p < .*01), and adequate construct and predictive validity. The total score is a sum of all items (some reversed). Higher scores indicate more dysregulation. In the present study, Cronbach’s α for the total score at intake was .95. The correlation between screen and intake DERS total score for 133 participants was .65.

**Outcome Questionnaire-45 (OQ-45).** The OQ-45 (52) is a 45-item instrument that can capture psychopathology changes through treatments. It consists of subscales that identify three types of problems that lead to general stress: psychological symptoms, interpersonal conflicts, and problems with social roles (53). Items are rated on a Likert scale ranging from 0 (*never*) to 4 (*almost always*). The OQ-45 demonstrates adequate test-retest reliability (*ρ*_I_ = .84) and excellent internal consistency (*α* = .93) (53). At intake, Cronbach’s *α* for the total score was .95.

**The Work and Social Adjustment Scale (WSAS).** The WSAS (54) is a 5-item measure examining the social and functional impairment stemming from a specific problem. In this study, we defined this specific problem to be emotional dysregulation (e.g., “Because of my difficulties managing emotions, my ability to work is impaired”). The questions are rated on a 9-point Likert scale (0 = my problem does not affect this at all; 8 = my problem affects this very seriously), and a total score was computed by summing all items. A total WSAS score above 10 suggests clinical levels of impairment and the measure has evidence for strong test-retest reliability, criterion validity, and sensitivity to change (55). In this study at pretreatment, Cronbach’s alpha for the total score was .86.

**Subjective Units of Distress Scale (SUDS).** During the MRI sessions, neurostimulation session, at follow up, and during the ambulatory assessment, we asked participants to rate their current distress on a scale from 0 – no distress to 9 – extreme distress (56).

**Manipulation Check.** During the neurostimulation session and at the 1 month follow up we examined dissociation during each baseline and regulation period using a 4 item scale (57). At the end of the intervention and at the 1 week and 1 month follow up participants were asked to rate on a scale from 1 (I am certain I received sham stimulation) to 9 (I am certain I received active stimulation) their confidence in the assigned condition that they were kept blind to. A rating of 5 indicated uncertainty. We also asked after the intervention for participants to give their best guess whether they received real or sham neurostimulation (force choice question).

**Tolerability Questionnaire.** Before and after the intervention session, participants were asked to rate on a scale from 0–3 (absent, mild, moderate, severe) the intensity of their headache, neck pain, scalp pain, seizure (as observed by technician), hearing impairment and any other side effect that they might have experienced from the TMS treatment.

**Exit Interview.** An unpublished, previously developed in-house interview (58) was used to examine feasibility and acceptability as directly relevant to the study. The interview was administered at the 1-month follow up, and it included open-ended questions about the overall experience as well as Likert-type questions about feasibility of the intervention (e.g., difficulty with limiting movement, level of comfort, ability to concentrate given the TMS noise, distress about the procedures, ease to hear and understand clinician, connection with clinician, and session engagement), acceptability (of session length, skills training, TMS procedures, personalized stressors use, ambulatory phone assessment) and overall satisfaction (i.e., likelihood to recommend to someone else). Feasibility and acceptability questions were rated on a scale from 0 (not at all) to 9 (extremely) and scores were reversed as needed and averaged in order to compute an overall feasibility and acceptability score where 0 represented not feasible/acceptable at all and 9 represented very feasible/acceptable. Satisfaction was rated on a 0 (low) to 100 (high) continuous scale. Two participants had to discontinue the study early, because of changes in medication. For these participants, the exit interview was conducted at the 1 week follow up assessment, right before discontinuing the study.

### *Psychophysiological Data*

HF-HRV was calculated from psychophysiological data collected during the intervention and follow-up sessions. AcqKnowledge software was used to collect and pre-process raw ECG data by using a built in automated HRV analysis tool that follows established frequency domain algorithm guidelines (59). Prior to the analysis, the continuous raw ECG data was visually inspected and artifacts cleaned following established guidelines (60). Then the AcqKnowledge automated tool was used to detect R-waves in the cleaned ECG data and to create a series of inter-beat intervals that was converted to a continuous, time-domain representation of HR using cubic-spline interpolation, which was resampled at 8-Hz. Spectral analysis was used to extract frequency information from resampled intervals, which was displayed as a Power Spectral Density periodogram. Summary report values for HRV frequency bands, including the high-frequency band of interest in this study, were also provided. Each session period (e.g., baseline, habituation, stressor, regulation) was divided into 120-s bins, and HF-HRV was extracted from each bin. One hundred and twenty seconds were chosen because the emotion induction period was 120-s long, and segments were intended to be equivalent in length across the experiment. Therefore, two HF-HRV values for each baseline, one for each stressor, and five values for each regulation period and habituation were computed.

The ECG signal was transformed into beats per minute (heart rate; HR) using the ‘Find Rate’ function of AcqKnowledge, using a moving average with a window of 15 seconds. For each baseline (session baseline and three pre-stimulus baselines), HR was averaged from the last 240 of the total 300 s. We excluded the first 60 s from each baseline to allow participants to settle in and relax to have a good representation of the physiological baseline. In cases where there was a clear spike in HR (e.g., because the participant coughed, talked, moved abruptly, etc), this artifact was removed before the average HR was calculated.

**Statistical Analyses**

Analyses were conducted using SPSS version 25.0.To test immediate effects of the intervention, we conducted three analyses examining HF-HRV, SUDS, and regulation duration. The treatment condition (active left or sham), the experimental condition (regulation 1, 2, and 3), the time within each experimental condition (coded 0-4 for each 120s segment within that period), and baseline were used to predict HF-HRV. Baseline HF-HRV was measured at the beginning of the experiment (session baseline) and right before each autobiographical stressor presentation (pre-stimulus baseline). Both session and pre-stimulus baselines were included as covariates because active rTMS may have cumulative effects (61) that could influence the pre-stimulus baselines.

Treatment condition, time in the experiment (post-baseline, post habituation, post stressor 1, post regulation 1, etc.), and planned covariates were used to predict SUDS throughout the experiment. In addition, we conducted a MMANOVA examining regulation duration during each regulation period. Effect sizes for these models were computed either by subtracting the adjusted means and dividing the difference by the baseline SD for the measure or by using Feingold’s formula (62) and interpreted using (63) specifications. To compute an effect size for regulation duration, we extracted the standard deviation from the same variable in a previous study (58), S.D. = 0.47 over 91 observations of regulation duration. Time was considered a categorical variable.

To test the long-term effects of the intervention, six MMANOVA models were conducted For the longitudinal self-reports (DERS, OQ-45, WSAS, and ERQ Reappraisal), time was entered as a categorical variable and intake measurements were co-varied. HF-HRV was extracted from the first two 120s segments from the 300 s regulation period at follow up. These two time points were entered in the analysis as a categorical time variable and the treatment condition and baseline HF-HRV (extracted from the last 120s of baseline) were added as covariates. Treatment condition, time in the follow up stressor task (post-baseline, post stressor, post regulation), SUDS baseline (collected at the beginning of the 1 month follow up day) were used to predict SUDS at follow up.

**Missing Data**

Across the 11 self-report and physiological variables examined for those included, 5.06% of the data was missing. Two participants did not complete the OQ-45 and one did not complete the WSAS at intake because of an administrative error. One participant changed medications right after their intervention session, and therefore was withdrawn from the study and did not complete the 1 week follow up assessment. Another participant changed medications right at the 1 week follow up assessment and therefore was withdrawn from the study and did not complete the 1 month follow up assessment. Both of these participants did complete the exit interview before being withdrawn from the study. Three participants were lost to follow up by the 1-month assessment, and one participant completed the 1 month remotely and therefore did not go through the 1-month stressor task. The psychophysiological data from one participant at follow up was accidentally deleted (but self-report was recorded and included in the follow up analyses). At the intervention session, one participant had all the SUDs data erased by accident (except for session baseline and habituation). All available data for this participant, including the recorded psychophysiological data, were included in analyses.

Following some of the autobiographical stressors, the HR was not higher then baseline. Therefore, some regulation durations were marked as “not stressed” and not included in analyses. Specifically, 6 participants did not stress following stresor1, 8 did not stress after stressor 2, and 3 did not stress after stressor 3 during the intervention day. At the one month follow up, 13 participants did not stress following the stressor task.

With regards to ambulatory calls data, one participant completed only 2 days of calls and one did not complete any calls because they were withdrawn from the study. Of the 1512 recorded calls, 1267 (83.8%) were placed and in 16.2% of cases (245 calls) the participants called in. Valid data was obtained in 1128 calls (74.6% of the total calls).

References

1.	First MB, Williams JB, Karg RL. Structured Clinical Interview for DSM-5 Disorders (Research Version: SCID-5-RV). Washington, DC: American Psychiatric Publishing; 2015.

2.	Gratz KL, Roemer L. Multidimensional Assessment of Emotion Regulation and Dysregulation: Development, Factor Structure, and Initial Validation of the Difficulties in Emotion Regulation Scale. J Psychopathol Behav Assess. 2004;26(1):41-54.

3.	Neacsiu AD, Eberle JW, Kramer R, Wiesmann T, Linehan MM. Dialectical behavior therapy skills for transdiagnostic emotion dysregulation: A pilot randomized controlled trial. Behav Res Ther. 2014;59:40-51.

4.	Whiteside U, Chen E, Neighbors C, Hunter D, Lo T, Larimer M. Difficulties regulating emotions: Do binge eaters have fewer strategies to modulate and tolerate negative affect? Eating Behaviors. 2007;8(2):162-9.

5.	Salters-Pedneault K, Roemer L, Tull MT, Rucker L, Mennin DS. Evidence of Broad Deficits in Emotion Regulation Associated with Chronic Worry and Generalized Anxiety Disorder. Cogn Ther Res. 2006;30(4):469-80.

6.	Fox HC, Axelrod SR, Paliwal P, Sleeper J, Sinha R. Difficulties in emotion regulation and impulse control during cocaine abstinence. Drug and Alcohol Dependence. 2007;89(2–3):298-301.

7.	Harrison A, Sullivan S, Tchanturia K, Treasure J. Emotion recognition and regulation in anorexia nervosa. Clin Psychol Psychother. 2009;16(4):348-56.

8.	Cohn AM, Jakupcak M, Seibert LA, Hildebrandt TB, Zeichner A. The role of emotion dysregulation in the association between men’s restrictive emotionality and use of physical aggression. Psychol Men Masc. 2010;11(1):53.

9.	Tull MT, Barrett HM, McMillan ES, Roemer L. A preliminary investigation of the relationship between emotion regulation difficulties and posttraumatic stress symptoms. Behavior Therapy. 2007;38(3):303-13.

10.	Osborne TL, Michonski J, Sayrs J, Welch SS, Anderson LK. Factor structure of the difficulties in emotion regulation scale (DERS) in adult outpatients receiving dialectical behavior therapy (DBT). J Psychopathol Behav Assess. 2017;39(2):355-71.

11.	Tull MT, Gratz KL, McDermott MJ, Bordieri MJ, Daughters SB, Lejuez CW. The role of emotion regulation difficulties in the relation between PTSD symptoms and the learned association between trauma-related and cocaine cues. Subst Use Misuse. 2016;51(10):1318-29.

12.	Contardi A, Imperatori C, Penzo I, Del Gatto C, Farina B. The association among difficulties in emotion regulation, hostility, and empathy in a sample of young Italian adults. Front Psychol. 2016;7:1068.

13.	Helbig‐Lang S, Rusch S, Lincoln TM. Emotion regulation difficulties in social anxiety disorder and their specific contributions to anxious responding. Journal of clinical psychology. 2015;71(3):241-9.

14.	Allan NP, Norr AM, Macatee RJ, Gajewska A, Schmidt NB. Interactive effects of anxiety sensitivity and emotion regulation on anxiety symptoms. J Psychopathol Behav Assess. 2015;37(4):663-72.

15.	Gross JJ, John OP. Individual differences in two emotion regulation processes: Implications for affect, relationships, and well-being. Journal of Personality and Social Psychology. 2003;85(2):348-62.

16.	Aldao A, Nolen-Hoeksema S, Schweizer S. Emotion-regulation strategies across psychopathology: A meta-analytic review. Clinical Psychology Review. 2010;30(2):217-37.

17.	Amstadter A. Emotion regulation and anxiety disorders. Journal of anxiety disorders. 2008;22(2):211-21.

18.	Melka SE, Lancaster SL, Bryant AR, Rodriguez BF. Confirmatory factor and measurement invariance analyses of the emotion regulation questionnaire. Journal of clinical psychology. 2011;67(12):1283-93.

19.	Salsman NL, Linehan MM. An investigation of the relationships among negative affect, difficulties in emotion regulation, and features of borderline personality disorder. Journal of Psychopathology and Behavioral Assessment. 2012;34(2):260-7.

20.	Dennis TA. Interactions between emotion regulation strategies and affective style: Implications for trait anxiety versus depressed mood. Motiv Emot. 2007;31(3):200-7.

21.	Fresco DM, Moore MT, van Dulmen MH, Segal ZV, Ma SH, Teasdale JD, et al. Initial psychometric properties of the experiences questionnaire: validation of a self-report measure of decentering. Behavior Therapy. 2007;38(3):234-46.

22.	Gillanders S, Wild M, Deighan C, Gillanders D. Emotion regulation, affect, psychosocial functioning, and well-being in hemodialysis patients. Am J Kidney Dis. 2008;51(4):651-62.

23.	Matsumoto D, Yoo SH, Nakagawa S. Culture, emotion regulation, and adjustment. Journal of personality and social psychology. 2008;94(6):925.

24.	Haga SM, Kraft P, Corby E-K. Emotion regulation: Antecedents and well-being outcomes of cognitive reappraisal and expressive suppression in cross-cultural samples. Journal of Happiness Studies. 2009;10(3):271-91.

25.	Zuurbier LA, Nikolova YS, Åhs F, Hariri AR. Uncinate fasciculus fractional anisotropy correlates with typical use of reappraisal in women but not men. Emotion. 2013;13(3):385.

26.	Vanderhasselt M-A, Baeken C, Van Schuerbeek P, Luypaert R, De Raedt R. Inter-individual differences in the habitual use of cognitive reappraisal and expressive suppression are associated with variations in prefrontal cognitive control for emotional information: an event related fMRI study. Biological Psychology. 2013;92(3):433-9.

27.	Swart M, Kortekaas R, Aleman A. Dealing with feelings: characterization of trait alexithymia on emotion regulation strategies and cognitive-emotional processing. PLoS One. 2009;4(6):e5751.

28.	Li Z, Sang Z, Zhang Z. Expressive suppression and financial risk taking: A mediated moderation model. Personality and Individual Differences. 2015;72:35-40.

29.	Egloff B, Schmukle SC, Burns LR, Schwerdtfeger A. Spontaneous emotion regulation during evaluated speaking tasks: associations with negative affect, anxiety expression, memory, and physiological responding. Emotion. 2006;6(3):356.

30.	Orgeta V. Avoiding Threat in Late Adulthood: Testing Two Life Span Theories of Emotion. Experimental Aging Research. 2011;37(4):449-72.

31.	Joormann J, Gotlib IH. Emotion regulation in depression: Relation to cognitive inhibition. Cognition and Emotion. 2010;24(2):281-98.

32.	Moore SA, Zoellner LA, Mollenholt N. Are expressive suppression and cognitive reappraisal associated with stress-related symptoms? Behaviour Research and Therapy. 2008;46(9):993-1000.

33.	Dunn LM. PPVT-revised manual. Circle Pines, MN: American Guidance Service; 1981 1981.

34.	Pitman RK, Orr SP, Forgue DF, de Jong JB, Claiborn JM. PSychophysiologic assessment of posttraumatic stress disorder imagery in vietnam combat veterans. Arch Gen Psychiatry. 1987;44(11):970-5.

35.	Schmahl CG, Elzinga BM, Ebner UW, Simms T, Sanislow C, Vermetten E, et al. Psychophysiological reactivity to traumatic and abandonment scripts in borderline personality and posttraumatic stress disorders: a preliminary report. Psychiatry Res. 2004;126(1):33-42.

36.	Esteban O, Markiewicz CJ, Blair RW, Moodie CA, Isik AI, Erramuzpe A, et al. fMRIPrep: a robust preprocessing pipeline for functional MRI. Nature methods. 2019;16(1):111.

37.	Tustison NJ, Avants BB, Cook PA, Zheng Y, Egan A, Yushkevich PA, et al. N4ITK: improved N3 bias correction. IEEE Trans Med Imaging. 2010;29(6):1310-20.

38.	Fonov VS, Evans AC, McKinstry RC, Almli C, Collins D. Unbiased nonlinear average age-appropriate brain templates from birth to adulthood. NeuroImage. 2009(47):S102.

39.	Pruim RH, Mennes M, van Rooij D, Llera A, Buitelaar JK, Beckmann CF. ICA-AROMA: A robust ICA-based strategy for removing motion artifacts from fMRI data. NeuroImage. 2015;112:267-77.

40.	Smith SM. Fast robust automated brain extraction. Hum Brain Mapp. 2002;17(3):143-55.

41.	Katz L, Epstein S. Constructive thinking and coping with laboratory-induced stress. Journal of Personality and Social Psychology. 1991;61(5):789.

42.	Beck A, Emery G, Greenberg R. Anxiety Disorders and Phobias: A Cognitive Perspective Basic Books. New York. 1985.

43.	Lazarus RS, Folkman S. Stress, appraisal, and coping: Springer publishing company; 1984.

44.	Meichenbaum D. Stress inoculation training: Pergamon; 1985.

45.	Beck AT, Dozois DJ. Cognitive therapy: current status and future directions. Annu Rev Med. 2011;62:397-409.

46.	Powers JP, LaBar KS. Regulating emotion through distancing: A taxonomy, neurocognitive model, and supporting meta-analysis. Neuroscience and biobehavioral reviews. 2019;96:155-73.

47.	Shurick AA, Hamilton JR, Harris LT, Roy AK, Gross JJ, Phelps EA. Durable effects of cognitive restructuring on conditioned fear. Emotion. 2012;12(6):1393-7.

48.	Ochsner KN, Bunge SA, Gross JJ, Gabrieli JDE. Rethinking feelings: an FMRI study of the cognitive regulation of emotion. J Cogn Neurosci. 2002;14(8):1215-29.

49.	Kendall PC. Coping cat workbook: Workbook Pub; 2006.

50.	First MB, Williams JB, Benjamin LS, Spitzer RL. SCID-5-PD: Structured clinical interview for DSM-5® personality disorders: American Psychiatric Association Publishing; 2016.

51.	Ventura J, Liberman RP, Green MF, Shaner A, Mintz J. Training and quality assurance with the Structured Clinical Interview for DSM-IV (SCID-I/P). Psychiatry Research. 1998;79(2):163-73.

52.	Lambert MJ, Burlingame GM, Umphress V, Hansen NB, Vermeersch DA, Clouse GC, et al. The reliability and validity of the Outcome Questionnaire. Clinical Psychology & Psychotherapy: An International Journal of Theory and Practice. 1996;3(4):249-58.

53.	Wells MG, Burlingame GM, Lambert MJ, Hoag MJ, Hope CA. Conceptualization and measurement of patient change during psychotherapy: Development of the Outcome Questionnaire and Youth Outcome Questionnaire. Psychotherapy: Theory, Research, Practice, Training. 1996;33(2):275.

54.	Mundt JC, Marks IM, Shear MK, Greist JM. The Work and Social Adjustment Scale: a simple measure of impairment in functioning. The British Journal of Psychiatry. 2002;180(5):461-4.

55.	Pedersen G, Kvarstein E, Wilberg T. The Work and Social Adjustment Scale: Psychometric properties and validity among males and females, and outpatients with and without personality disorders. Personality and mental health. 2017;11(4):215-28.

56.	Wolpe J. The practice of behavior therapy. New York, NY <etc.>: Pergamon Press; 1969.

57.	Stiglmayr C, Schmahl C, Bremner JD, Bohus M, Ebner-Priemer U. Development and psychometric characteristics of the DSS-4 as a short instrument to assess dissociative experience during neuropsychological experiments. Psychopathology. 2009;42(6):370-4.

58.	Neacsiu AD, Beynel L, Powers JP, Szabo ST, Appelbaum LG, Lisanby SH, et al. Enhancing Cognitive Restructuring with Concurrent Repetitive Transcranial Magnetic Stimulation for Transdiagnostic Psychopathology: A Proof of Concept Randomized Controlled Trial. medRxiv. 2021:2021.01.18.21250060.

59.	Camm A, Malik M, Bigger J, Breithardt G, Cerutti S, Cohen R, et al. Heart rate variability: standards of measurement, physiological interpretation and clinical use. Task Force of the European Society of Cardiology and the North American Society of Pacing and Electrophysiology. Circulation. 1996;93(5):1043-65.

60.	Berntson GG, Bigger JT, Jr., Eckberg DL, Grossman P, Kaufmann PG, Malik M, et al. Heart rate variability: origins, methods, and interpretive caveats. Psychophysiology. 1997;34(6):623-48.

61.	Bäumer T, Lange R, Liepert J, Weiller C, Siebner HR, Rothwell JC, et al. Repeated premotor rTMS leads to cumulative plastic changes of motor cortex excitability in humans. Neuroimage. 2003;20(1):550-60.

62.	Feingold A. Effect sizes for growth-modeling analysis for controlled clinical trials in the same metric as for classical analysis. Psychol Methods. 2009;14(1):43.

63.	Cohen J. Statistical power analysis for the behavioral sciences Rev. ed. ed. New York, NY: Academic Press; 1977.
